# “Corticosteroid pulses for hospitalized patients with COVID-19: Effects on mortality”

**DOI:** 10.1101/2020.09.30.20204719

**Authors:** Ivan Cusacovich, Álvaro Aparisi, Miguel Marcos, Cristina Ybarra-Falcón, Carolina Iglesias-Echevarria, Maria Lopez-Veloso, Julio Barraza-Vengoechea, Carlos Dueñas, Santiago Antonio Juarros Martínez, Beatriz Rodríguez-Alonso, José-Ángel Martín-Oterino, Miguel Montero-Baladia, Leticia Moralejo, David Andaluz-Ojeda, Roberto Gonzalez-Fuentes

**Author notes:** Corresponding author: Ivan Cusacovich, Internal Medicine Department. Hospital Clínico Universitario de Valladolid, Spain., Ramón y Cajal 3. 47005. Valladolid. Spain. These authors contributed equally (David Andaluz-Ojeda, Roberto Gonzalez-Fuentes). Email (in order of appearance). Conflicts of interest: None. Financial sources: None. Authors’ contributions: All authors contributed to data collection. Ivan Cusacovich, Álvaro Aparisi, Miguel Marcos David Andaluz-Ojeda, Roberto Gonzalez-Fuentes, and Carlos Dueñas contributed to the study design. Ivan Cusacovich and David Andaluz-Ojeda performed the statical analysis. Ivan Cusacovich, Alvaro Aparisi, David Andaluz-Ojeda, Roberto Gonzalez-Fuentes, Miguel Marcos, and Carlos Dueñas contributed to writting the draft. Ethics committee approval: Approved by the local ethical committee (CEIC) ID. Number: PI 20-1812-COVID.

## Abstract

Background: COVID-19 has high mortality in hospitalized patients, and we need effective treatments. Our objective was to assess corticosteroid pulses’ influence on 60-days mortality in hospitalized patients with severe COVID-19, intensive care admission, and hospital stay. Methods: We designed a multicenter retrospective cohort study in three teaching hospitals of Castilla y León, Spain (865.096 people). We selected patients with confirmed COVID-19 and lung involvement with a pO2/FiO2 < 300, excluding those exposed to immunosuppressors before or during hospitalization, patients terminally ill at admission, or died the first 24 hours. We performed a propensity score matching (PSM) adjusting covariates that modify the probability of being treated. Then we used a Cox regression model in the PSM group to consider factors affecting mortality. Findings: From 2933 patients, 257 fulfilled the inclusion and exclusion criteria. One hundred and twenty-four patients were on corticosteroid pulses, and 133 were not. 30·3% (37/122) of patients died in the corticosteroid pulses group and 42·9% (57/133) in the non-exposed cohort. These differences (12·6% CI95% [8·54-16·65]) were statically significant (log-rank 4·72, p=0·03). We performed PSM using the exact method. Mortality differences remained in the PSM group (log-rank 5·31, p=0·021) and were still significant after a Cox regression model (HR for corticosteroid pulses 0·561, p= 0·039). There were no significant differences in intensive care admission rate (p=0·173). The hospital stay was longer in the corticosteroid group (p<0,001). Interpretation: This study provides evidence about treatment with corticosteroid pulses in severe COVID-19 that might significantly reduce mortality. Strict inclusion and exclusion criteria with that selection process set a reliable frame to compare mortality in both exposed and non-exposed groups. Funding: There was no funding provided.

## MAIN TEXT

### INTRODUCTION

In December 2019, a new betacoronavirus called SARS-Cov-2 induced severe bilateral pneumonia similar to severe acute respiratory syndrome (SARS), described in 2003. This coronavirus disease (COVID-19) had lower mortality than SARS-Cov-1 infection, but higher infective capacity. The epidemic began in Wuhan, mainland China, but in a few months became pandemic.

Spain was one of the most affected countries in the world, especially in Madrid, Catalonia, and Castilla y León regions ^(1)^.

After 32 885 641 confirmed cases, mortality rates are between 3-4%, mostly due to acute respiratory distress syndrome (ARDS) and micro-pulmonary embolism. These symptoms are related to a hyperinflammatory state and a cytokine storm syndrome in some patients ^(2)^. Thus, several authors have postulated that immunosuppressor agents (like corticosteroids, anakinra^(3)^ or tocilizumab ^(4)^) might be useful for these patients.

Several studies have tried corticosteroids for the treatment of viral pneumonia (including Flu and SARS-Cov1) and ARDS, with different results ^5-10^ Only a few studies demonstrate the benefits of corticosteroids on mortality ^11-14^. Preliminary results of the Recovery trial obtained mortality benefits with dexamethasone treatment in COVID-19 patients that required oxygen supplementation ^(15)^.

Corticosteroids inhibit the migration of leukocytes to inflamed tissues, enhancing their migration from bone marrow to blood ^(16)^ and decreasing leukocyte apoptosis ^(17)^. They also inhibit leukocyte reactive oxygen species, increase IL-10 ^(18),(19),^ and alter maturation and differentiation of dendritic cells ^(20),(21)^. Corticosteroids modify NK cytolytic activity and monocyte activation ^(21)^. They also downregulate IL-1, IL-2, IL-6, IL-8, IFN-γ, or TNF-α by transrepression ^(22)^.

The dose and the timing of corticosteroids are essential to determine their effect. There are three moments in which the use of corticosteroids might be especially useful. These are the onset of acute lung injury, the initial phase of ARDS, and ARDS refractory to treatment ^(23)^.

At thirty to one hundred mg of prednisone equivalent daily dose, corticosteroids act over cytosolic glucocorticoid receptors (cGCR), following the so-called genomic pathway ^(22),(24)^. The genomic pathway effect is highest at 100 mg. The complex formed by glucocorticoid and its cytosolic GCR has two actions: Promotion of anti-inflammatory transcription factors (transactivation) like IL-10 and annexin 1, and inhibition of inflammatory transcription factors (transrepression) like IL-1, IL-2, IL-6, interferon-γ (IFN—γ), prostaglandins or tumor necrosis factor α (TNF-α), and IL-8. All these changes carry out from hours to days.

If we use an equivalent dose of prednisone higher than 100 mg daily (so-called pulse corticosteroids), we obtain the maximum effect of the genomic pathway, and additional responses from the faster “non-genomic pathway” ^(22)^. These non-genomic mechanisms include membrane dysfunction in all immune cells (including lymphocytes), with a delayed flow across the membrane in the calcium and sodium channels with subsequent decreased ATP production. Other non-genomic effects bind to membrane GCR in T cells ^(22)^ or Src protein release from the complex cGCR-multiprotein (anti-inflammatory effects). This quick (in hours) and effective action^(25)^ justify their use in life-threatening situations in autoimmune diseases.

### METHODS

We analyzed patients with COVID-19 admitted between Mar 12th and May 20th to three tertiary teaching hospitals in Castilla y León, Spain: Hospital Clínico Universitario de Valladolid (HCUV), Hospital Universitario de Salamanca (HUSA), and Hospital Universitario de Burgos (HUBU). The three hospitals cover all hospital admissions in a geographical area corresponding to 865 096 people.

The treating team decided the prescription of all drugs, without any intervention from investigators. We obtained the local ethics committee (CEIC) permission to perform the study. Informed consent was obtained. We designed a retrospective cohort study and compared a cohort of patients exposed to corticosteroid pulses and an unexposed one.

#### Data source

We analyzed paper and electronic records in all hospitals. We recorded variables related to clinical outcomes and corticosteroids exposure (supplementary material).

#### Inclusion and exclusion criteria

We included patients older than 18 years, testing positive on SARS-Cov-2 PCR (nasopharyngeal or oropharyngeal swab specimens). Patients with positive Elisa serology and consistent clinical symptoms were also considered confirmed cases.

All included patients had a significant lung involvement, defined as a pO2/FiO2 < 300, maintained for 24 hours or repeated for three days. We measured pO2/FiO2 in arterial gasometry or estimated it from pulse oximetry data (non-linear estimate model) (26).

We excluded patients receiving classic immunosuppressors or cytokine blockers (as cyclosporine, tocilizumab, or anakinra). Concomitant drugs allowed were hydroxychloroquine, azithromycin, remdesivir, lopinavir/ritonavir, and colchicine. We excluded patients who died in the first 24 hours of admission. Patients on corticosteroid treatment in a different regimen than the one described in this study were excluded. We also excluded pregnant women, terminally ill patients, and patients under a limitation of therapeutic efforts during the first 24 hours of admission.

#### Corticosteroids pulses definition

We considered exposure to corticosteroid pulses if administered at a daily dose of 125 to 500 mg of intravenous methylprednisolone from two to five days. We did not include patients with repeated corticosteroid pulses nor treatments longer than five days. About timing, we considered corticosteroid pulses in the ± 3 days, respecting the inclusion criterion date.

#### Endpoints

The primary endpoint was 60-days mortality in exposed versus non-exposed patients. Secondary endpoints were 30-days mortality, intensive care unit (ICU) admission, in-hospital stay, viral shedding until negative PCR, and serious adverse events, including infections.

#### Statical Analysis

We expressed continuous variables with the median and interquartile range (mean and standard deviation if they had normal distribution). We used Chi-square to compare qualitative variables and the T-test (if normal distribution) or the Mann Withney test to compare two quantitative variables. We performed the Kolmogorov-Smirnov test to examine normal distribution.

We performed a propensity score matching to balance the difference of covariates related to exposure to corticosteroid pulses.

To select suitable covariates to control, we set biologically plausible variables related to the probability of being treated with corticosteroids pulses (propensity score). First, we analyzed variables associated with the propensity score in univariate analysis. Variables found significant were dichotomized, and then we performed a binary logistic regression to evaluate independent variables associated with the propensity score. We chose three matching methods (propensity score matching) to preprocess the sample: Nearest neighbor, the nearest neighbor with a caliper (at a distance of 0·05, 0·1, 0·2, and 0·3), and exact matching. We performed the propensity score matching using the R software, with the MatchIt and Cobalt libraries.

We checked the balance of the propensity score matching though the “difference of means” to ensure that the covariates’ distribution was similar in the treated and control group, and we picked the best-matched model.

Once we completed the propensity score matching, we performed a Cox proportional hazard regression analysis to evaluate mortality and intensive care admission.

### RESULTS

#### Patients

From 2933 patients in our cohort, 257 fulfilled the inclusion and exclusion criteria. 767 fulfilled the inclusion criterion, and 546 had any exclusion criteria (See Figure 1). We diagnosed with COVID-19 in 243 patients based on SARS-Cov-2 PCR in the nasopharyngeal or oropharyngeal swabbing and 14 patients based on positive serology with compatible symptoms. 124 patients were on corticosteroid pulses, and 133 were not.

**Figure-1.**
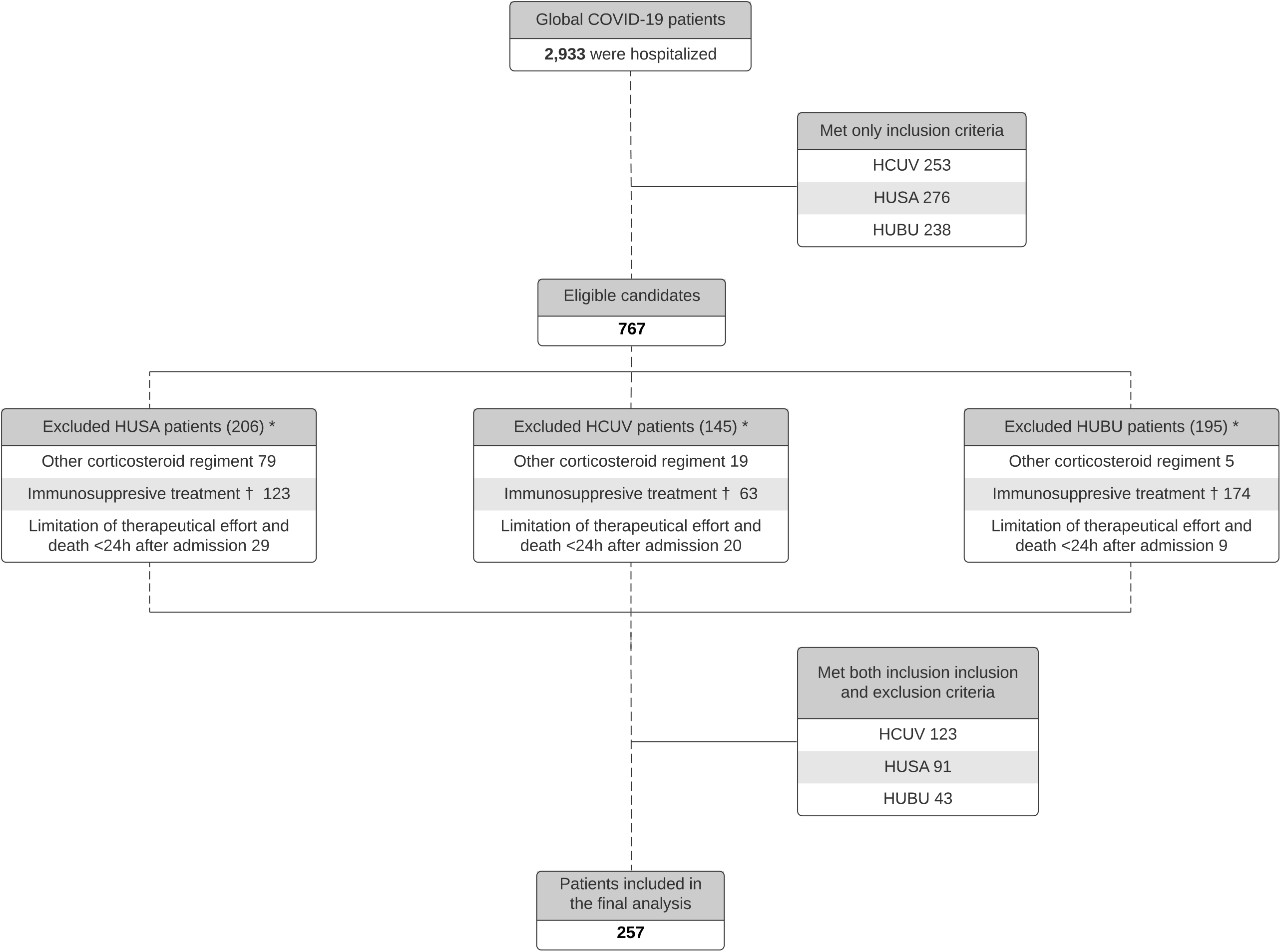
Flow Diagram of COVID-19 included patients in this study.

#### Propensity score matching

We calculated the propensity score (probability of being treated with corticosteroid pulses) in each participant from a binary logistic regression. Variables statically significant in the binary logistic regression were: Epidemiological week, presence of bilateral infiltrates or not, Center in Castilla y León, Ferritin, and COVID gram score (see supplementary material).

After performing the propensity score, we tried several preprocessing methods for matching (see Methods), and we selected the exact matching method as it got the minimum difference between groups with the minimum sample loss (28 controls and 2 treated patients).

The “difference of means” was zero in the treated versus the controls because we used the exact matching method (Figure 2 and supplementary material).

**Figure-2.**
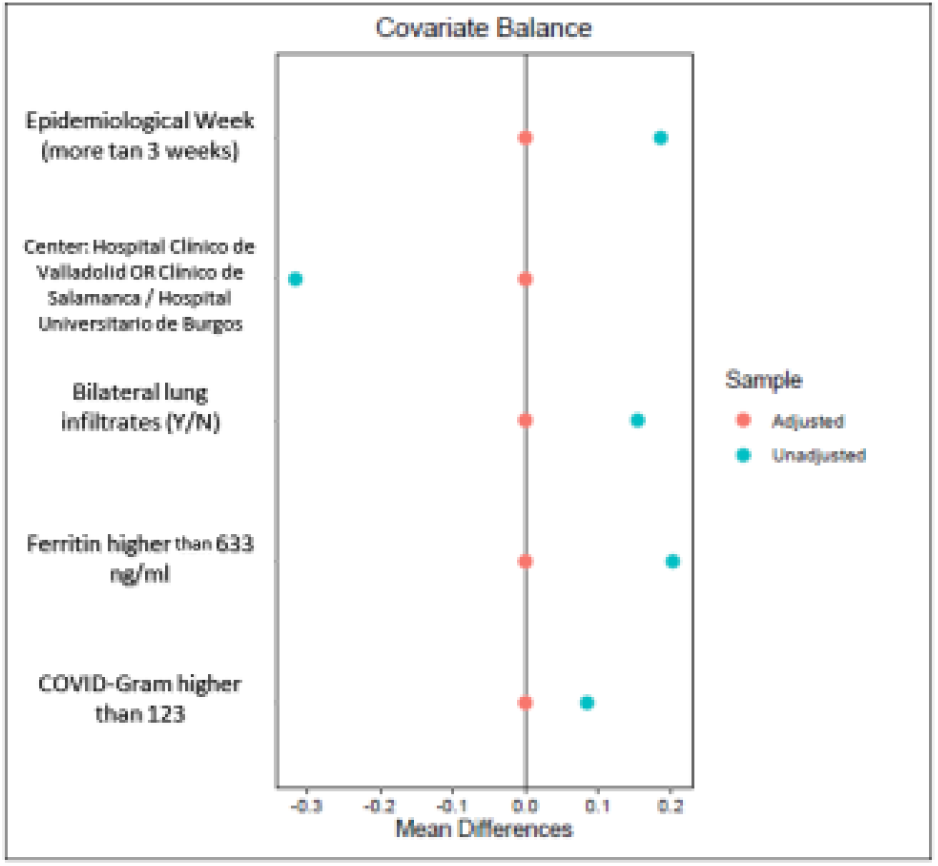
Love Plot of the propensity score matching using the Exact Method

After the propensity score matching, the sample consisted of 207 patients (119 treated and 88 controls).

#### Baseline features

The median Age in all participants (257) was 75 [63·5-83] years. One hundred and eleven participants (43·2%) were women. Their median classic Charlson score was 1 [0-3].

Comparing patients exposed to corticosteroid pulses and the non exposed ones, we found that Age and comorbidities were similar in both groups, without significant differences. The COVID-gram score (27) was 155.8 in the bolus group and 152·3 in the control group, even decreasing these differences after matching. The classic Charlson score was significantly higher (0·7 points) in the control group (p=0·012). These differences disappeared after dichotomizing the variable in the matched group (≤2 or >2 comorbidities, p= 0·171). (See table 1)

**Table-1.** Baseline features of patients from all centers combined

| Treatment | Overall population |  | Matched population |  |
| --- | --- | --- | --- | --- |
|  | Methylprednisolone<br>(n=124) | Usual care<br>(n=133) | Methylprednisolone<br>(n=117) | Usual care<br>(n=88) |
| **Baseline characteristics** |  |  |  |  |
| Gender (male) no- (%) | 77 (62%) | 69 (51.8%) | 74 (63.3%) | 45 (51.1%) |
| Age (mean-years) | 74 [59-83] | 76 [65-84] | 75[60-83] | 76 [66-83] |
| > 80 years no- (%) | 37 (29.8%) | 47 (35.3%) | 36 (30.8%) | 36 (30.8%) |
| Autoimmune disease | 4 (3.2%) | 3 (2.3%) | 3 (2.5%) | 3 (3.4%) |
| Cancer (%) | 17 (13.7%) | 17 (12.8%) | 17 (14.5%) | 11 (12.5%) |
| Chronic Kidney disease (%) | 6 (4.8%) | 14 (10.5%) | 6 (5.1%) | 9 (10.2%) |
| Dementia (%) * | 7 (5.6%) | 21 (18.7%) | 6 (5.1%) | 12 (13.6%) |
| Diabetes | 40 (32.3%) | 52 (39.1%) | 38 (32.5%) | 36 (40.9%) |
| Dyslipemia | 44 (35.8%) | 40 (30.1%) | 42 (35.9%) | 29 (33%) |
| Hypertension | 63 (50.8%) | 67 (50.4%) | 42 (35.9%) | 50 (56.8%) |
| Obesity | 22 (17.8%) | 18 (13.5%) | 20 (17.1%) | 13 (14.7%) |
| Smoker | 32 (25.8%) | 29(21.8%) | 30(25.6%) | 15 (17%) |
| Prior AIT/Stroke (%) | 7 (5.6%) | 9 (6.8%) | 7 (6%) | 5 (5.7%) |
| Prior IHD (%) | 12 (9.7%) | 12 (9%) | 12 (10.2%) | 10 (11.4%) |
| Prior lung disease | 20 (16.1%) | 23 (17.3%) | 20 (17.1%) | 17 (19.3%) |
| Charlson comorbidity index | 1 [0-2] | 1 [0-4] | 1 [0-2] | 1 [0-2] |
| **Previous treatment** |  |  |  |  |
| Corticosteroids | 4 (3.2%) | 2 (1.5%) | 4 (3.4%) | 1 (1.2%) |
| Anticoagulants | 14 (11.2%) | 10 (7.5%) | 14 (11.9%) | 6 (6.8%) |
| **Main findings at admission** |  |  |  |  |
| Respiratory insufficiency | 104 (83.9%) | 105 (80.8%) | 99 (84.6%) | 73 (83.9%) |
| Bilateral infiltrates Chest X-ray | 103 (83.1%) | 91 (68.4%) | 105 (89.7%) | 67 (76.1%) |
| pO2/FiO2 | 239,5 [142,2-276,1] | 238,1[192.8-285.7] | 250 [153.2-276.2] | 238 [180,5-276,1] |
| COVID GRAM | 155.8 [83.3-230] | 152.3 [69-235.6] | 157.5 [83.6-231.6] | 157,7 [82-233,2] |
| **Laboratory findings at admission** |  |  |  |  |
| Glucose (mg/dl) | 114.5 [94.2-145] | 118.5 [100.1-168.2] | 114.6 [94.5-147] | 114.3 [99.2-180] |
| C-Reactive protein (mg/L) | 126.2 [84-200.2] | 85.2 [36.1-180] | 124.5 [66.7-194.5] | 111.5 [60.9-213.5] |
| Creatinine (mg/dL) | 1.06 [0.81-1.47] | 0.99 [0.78-1.53] | 1.06 [0.8-1.45] | 1.01 [0.79-1.62] |
| D-Dimer (ng/mL)* | 842 [448-1,450] | 1,105 [540,2-2,532] | 848 [4,127-1,425] | 1,250 [678-2,532] |
| Ferritin (ng/mL) | 1,533.5 [764-2,351.2] | 921 [359.5-1,466] | 1,475 [742-2,405] | 1,104 [556-1,650] |
| Interleukin-6 (pg/mL) | 35.5 [10.15-118.7] | 51[17.4-119] | 35.5 [10.1-118.7] | 59 [17.6-123] |
| Lactate dehydrogenase (UI/L) | 348,5 [283,7-460,7] | 312[248-394] | 341 [283-455] | 326 [259-396.5] |
| Lymphocytes (cells/mm<sup>3</sup>) | 1,000 [672-1,357] | 1,000 [715-1,415] | 1,000 [670-1,320] | 1,010 [730-1,467] |
| Neutrophils (cells/mm<sup>3</sup>x10<sup>3</sup>) | 5,050 [3,575-7,212] | 5,670 [3,895-8,805] | 5,997 [3,580-7,085] | 5,155 [133-179] |
| Procalcitonin (ng/ml) | 0.18 [0.1-0.37] | 0.15 [0.07-0.5] | 0.16 [0.1-0.3] | 0.19 [0.1-1.5] |
| **Specific COVID-19 treatment** |  |  |  |  |
| Azithromycin * | 115 (92.7%) | 94 (71.2%) | 108 (92.3%) | 64 (73.3%) |
| Interferon Beta-1b * | 48 (38.7%) | 23 (17.4%) | 44 (37.6%) | 20 (23%) |
| Hydroxychloroquine* | 120 (96.8%) | 121 (91.7%) | 113 (96.6%) | 80 (92%) |
| Lopinavir/ritonavir * | 106 (85.5%) | 92 (69.7%) | 102 (87.2%) | 65 (74.7%) |
| Colchicine * | 16 (12.9%) | 2 (1.5%) | 15 (12.8%) | 2 (2.3%) |
| **Non-specific COVID-19 treatment** |  |  |  |  |
| Prophylactic anticoagulation* | 69 (56.1%) | 90 (68.7%) | 66 (56.4%) | 57 (66.3%) |
| Intermediate anticoagulation* | 12 (9.8%) | 4 (3.1%) | 10 (8.5%) | 4 (4.7%) |
| Full anticoagulation * | 26 (21.1%) | 16 (12.2%) | 26 (22.2%) | 13 (15.1%) |

Data are shown as median (IQR) or n (%). (\*) Indicate a significant (P <0,05) difference between methylprednisolone and usual care population in both groups, the overall population, and propensity score match. It was calculated using the $\chi^2$ test or Kruskal-Wallis test as appropriate. PaO2/FiO2=ratio of arterial oxygen partial pressure to fractional inspired oxygen.

LDH and ferritin at admission were higher in the pulses group, but these differences disappeared after matching. Peak ferritin and peak LDH during hospitalization were significantly higher in the pulses group, even after matching (see table 1).

Concomitant treatments with colchicine, interferon beta-1b, lopinavir/ritonavir, and azithromycin were more common in the corticosteroid pulses group than in controls, both before and after matching (see table 1).

We did not find differences in pO2/FiO2 between the pulses and control group (p=0·183 in all participants and p=0·69 in the matched group), but bilateral lung infiltrates were more frequent in the corticosteroid pulses group (p=0.006). That difference disappeared after matching (p=0·299).

#### Outcomes

#### Primary endpoint

Ninety-four patients died during the 60 days after admission, representing 36·9 % (94/255) of the sample. 30·3% (37/122) of patients died in the corticosteroid pulses group and 42.9% (57/133) in the non-exposed cohort. These differences (12·6%, CI95% [8·54-16·65]) were statically significant in the Kaplan Meier curve (log-rank 4·72, p=0·03). (See figure 3)

**Figure-3.**
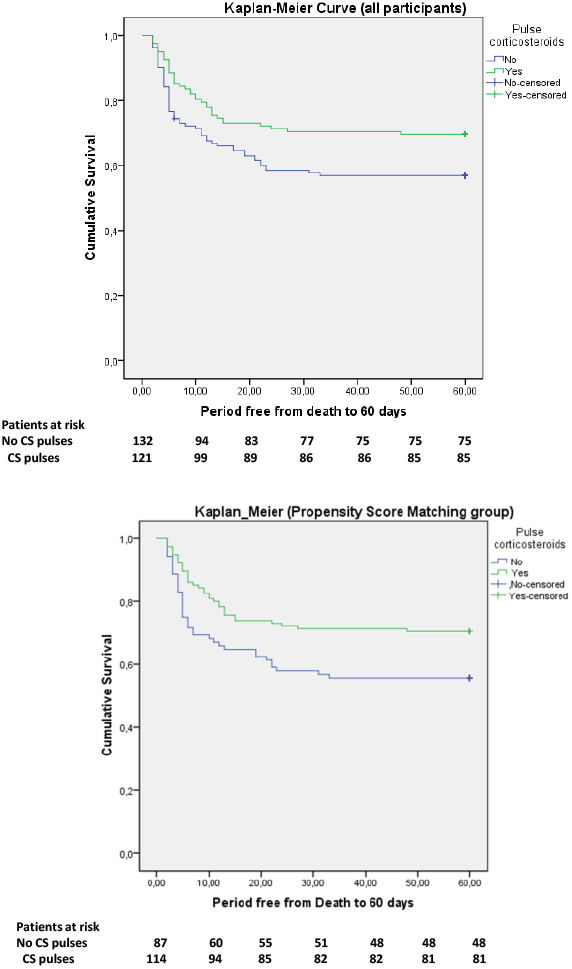
Kaplan-Meier Estimates of 60-days Mortality in patients with and without corticosteroid treatment

We carried out a propensity score matching and calculated the mortality in the matched group. The corticosteroid pulses group had a 60 days mortality of 29·6% (34/115), while mortality in the control group was 44.·3% (39/88). Again these differences were statically significant (log-rank 5·31, p=0·021). (See figure 3 and table 2).

**Table-2.**
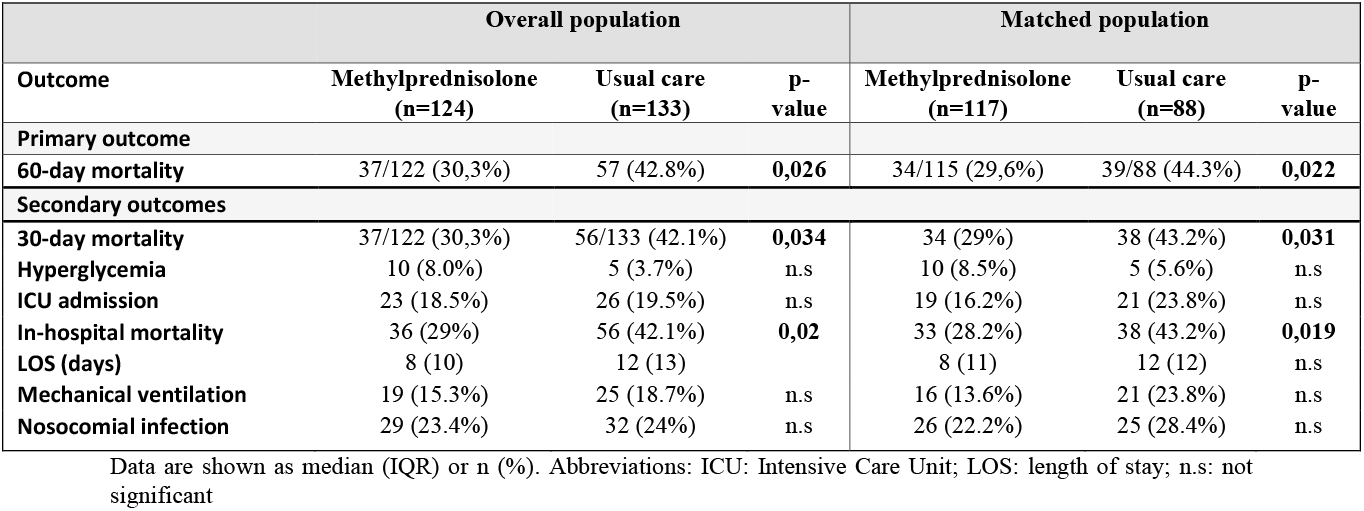
Primary and secondary outcomes of the global and matched population.

| Overall population |  |  |  | Matched population |  |  |
| --- | --- | --- | --- | --- | --- | --- |
| Outcome | Methylprednisolone<br>(n=124) | Usual care<br>(n=133) | p-value | Methylprednisolone<br>(n=117) | Usual care<br>(n=88) | p-value |
| **Primary outcome** |  |  |  |  |  |  |
| **60-day mortality** | 37/122 (30,3%) | 57 (42.8%) | **0,026** | 34/115 (29,6%) | 39/88 (44.3%) | **0,022** |
| **Secondary outcomes** |  |  |  |  |  |  |
| **30-day mortality** | 37/122 (30,3%) | 56/133 (42.1%) | **0,034** | 34 (29%) | 38 (43.2%) | **0,031** |
| **Hyperglycemia** | 10 (8.0%) | 5 (3.7%) | n.s | 10 (8.5%) | 5 (5.6%) | n.s |
| **ICU admission** | 23 (18.5%) | 26 (19.5%) | n.s | 19 (16.2%) | 21 (23.8%) | n.s |
| **In-hospital mortality** | 36 (29%) | 56 (42.1%) | **0,02** | 33 (28.2%) | 38 (43.2%) | **0,019** |
| **LOS (days)** | 8 (10) | 12 (13) |  | 8 (11) | 12 (12) | n.s |
| **Mechanical ventilation** | 19 (15.3%) | 25 (18.7%) | n.s | 16 (13.6%) | 21 (23.8%) | n.s |
| **Nosocomial infection** | 29 (23.4%) | 32 (24%) | n.s | 26 (22.2%) | 25 (28.4%) | n.s |

Data are shown as median (IQR) or n (%). Abbreviations: ICU: Intensive Care Unit; LOS: length of stay; n.s: not significant

We performed a multivariate analysis using a Cox regression model in the propensity score matching group, after dichotomizing variables, and those independently related to 60-days mortality were: Corticosteroid pulses (HR 0·561, p= 0·039), Age older than 80 years (HR 7·3, p<0001), CPR at admission higher than 200 mg/dl (HR 3·35, p<0,001), neutrophil/lymphocyte index >7·4 (HR 2·15, p= 0·010), Charlson index higher than 2 points (HR 2·15, p=0.·018), and LDH at admission >372 UI/ml (HR 2·29, p=0·008). (See table 4 and figure 4). In the equation, we did not include other variables (like pO2/FiO2, lung infiltrates, or D-Dimer) that were not significant in the multivariate model, although important in the univariate analysis.

**Figure-4.**
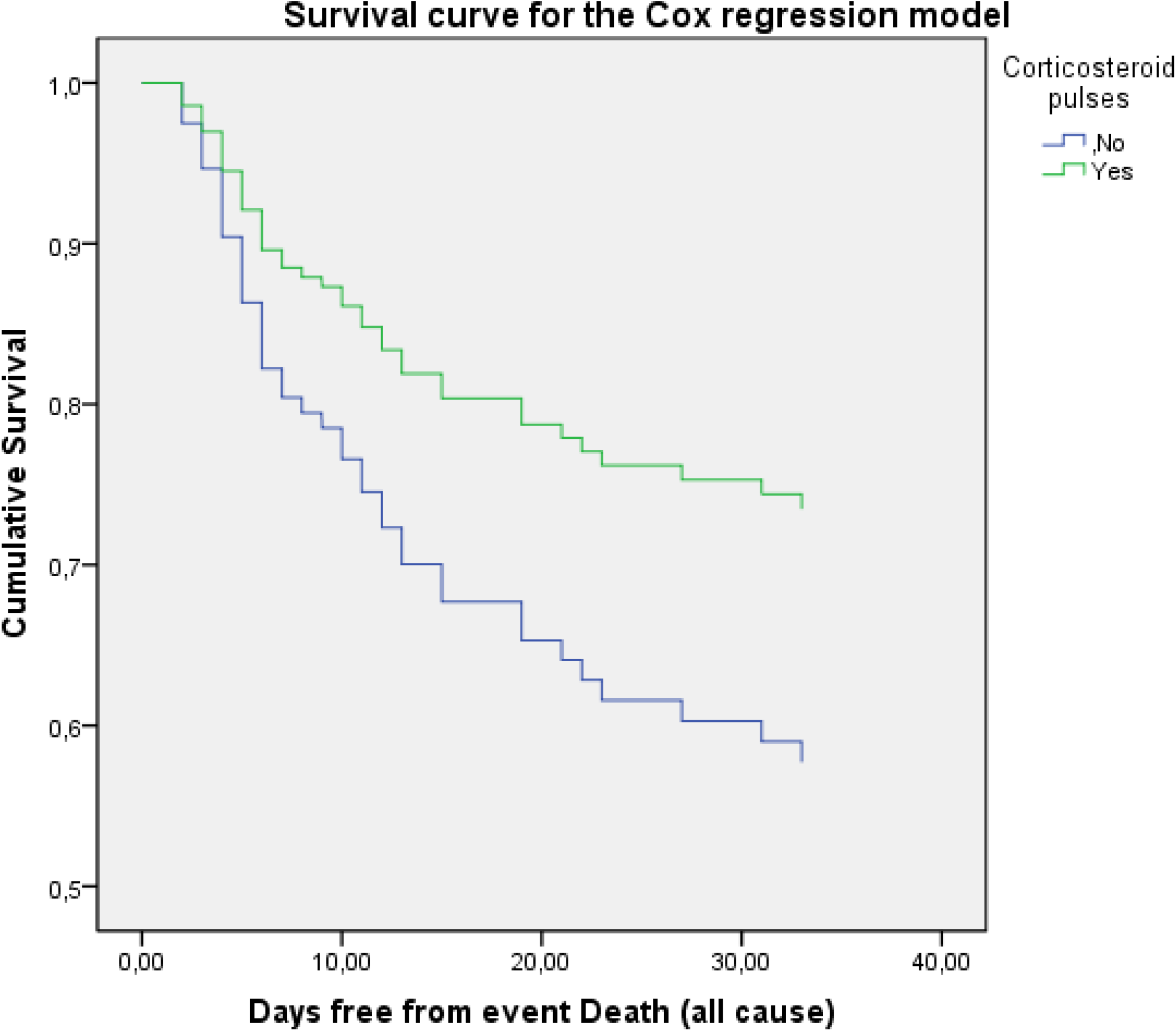
Survival for the Cox regression model in corticosteroids exposed and non-exposed patients (in the PSM group)

We studied the possible association of colchicine, azithromycin, and lopinavir/ritonavir on mortality, as those treatments were more frequently used in the corticosteroid pulses group. The three treatments were associated with lower mortality in the Kaplan-Meier curves (p=0·008 for colchicine, p=0·032 for lopinavir/ritonavir, and p<0·001 for azithromycin). These differences disappear after adjusting for Age higher than 80 years in both three drugs (p=0·1 for colchicine, p=0·794 for lopinavir/ritonavir, and p=0·378 for azithromycin).

#### Secondary endpoints

The thirty-days mortality was 30·3% (37/122) in the corticosteroid pulses group and 42·1% (56/133) in the non-exposed cohort. The difference was statically significant (log-rank=4·3, p=0·038) in the Kaplan-Meier curve. That difference remained in the propensity score matching group (p=0·03) (See supplementary material). After using the same Cox regression model used in the 60-days mortality analysis, corticosteroid pulses remain a protective factor for 30-days mortality (p=0·049). (See supplementary material and table 3).

**Table-3.** Cox regression for evaluating the 60-day mortality in the matched population

|  | Regression coefficient | HR (95% CI) | p-value |
| --- | --- | --- | --- |
| Corticosteroid pulses (Y/N) | -,578 | 0,561 (0,324-0,971) | 0,039 |
| Age more than 80 years | 1,995 | 7,355 (3,985-13,574) | <0,001 |
| CRP higher than 200 mg/dl | 1,211 | 3,358 (1,843-6,118) | <0,001 |
| Neutrophil / lymphocyte > 7.4 | ,873 | 2,394 (1,352-4,237) | 0,003 |
| Charlson index with ≥ 3 points | ,765 | 2,150 (1,203-3,844) | 0,010 |
| LDH at admission >372 UI/ml | ,786 | 2,194 (1,227-3,921) | 0,008 |

Forty-nine patients were admitted to ICU during this period. ICU admission was 18·5% (23/124) in the corticosteroid pulses cohort and 19.5% (26/133) in the non-exposed group (p=0·838). We found similar results in the propensity score matching group (ICU admission 16·2% in the corticosteroid group and 23·9% in the control group, p=0·173). The time from hospital admission to ICU admission was zero to four days, and 83·6% of patients were admitted in the first 24 hours. Mortality among the ICU admitted patients was 17·4% (4/23) in the exposed group and 61·5% (16/26) in the non-exposed group. This difference was statically significant for 60-days mortality (log-rank=9·7, p=0·002); This difference remains in the propensity score-matched group (15·8% vs. 61·9%, p=0·003). After adjusting for several variables (Peak LDH, Peak CRP, Number of comorbidities, D-dimer at admission, SaO2/FiO2, and Age) in a Cox regression, those differences in mortality were not significant (See supplementary material). The ICU average stay was 19·2 and 21·67 days for both exposed and non-exposed cohort, respectively (p=0·750).

The in-hospital median stay was 12 [7·25-19·75] days in the treated cohort and 8 [5-15] days in the non-exposed group (p<0·001). These differences remain in the matched group (p=0·001). However, differences were not statically significant if we analyzed them in the survivor’s group (difference of means 1·2 days, p=0·619).

Viral shedding until negative PCR was shorter (but not significant p=0·279) in the corticosteroid cohort (25·02 days in the corticosteroid group and 30·65 days in non-exposed).

We reported serious adverse events in 17 patients in the exposed cohort and 15 patients in the control group (p=0·133). Hemorrhage happened in 9 patients, without difference between groups (p=0·053). In-hospital infections were not higher in the corticosteroid bolus group (29/124) than in the non-exposed cohort (32/129), p=0·792.

We found 27 patients with pulmonary embolism by CT-scan (8 in the exposed group and 19 in non-exposed, p=0·066). We also reported two ischemic strokes and one acute myocardial infarction.

### DISCUSSION

Corticosteroid pulses have been widely used in Spain, especially in the Castilla y León region, but not in other countries for the treatment of COVID-19. The rationale of its use is to stop the systemic inflammation process ^(2)^ that develops in some patients with severe COVID-19. Some studies^(28),(29),(30)^ have described the positive effects of corticosteroid pulses on mortality in patients with severe COVID-19.

We found a significant improvement in survival in patients treated with corticosteroid pulses. We designed a retrospective cohort study to confirm this statement, which is the main limitation of our work. As there is no randomization, unknown confounders might be unattended.

We carried out a multicenter study with three teaching hospitals in the Castilla y León region in Spain. This fact is one of the main strengths of the study. On the one hand, we summarized various treatment protocols in each center, showing a wider specter of treatment options for severe COVID-19. This protocol variety determines different probabilities of being treated with corticosteroids in each center and enables us to adjust them in the latter propensity score matching. It also considers different hospital admission criteria and different extra-hospital resources that may change hospitalized patients’ baseline features.

On the other hand, it represents all hospital admissions in a geographic area in Castilla y León with more than 865 000 people that have similar epidemiological features. They also had the same timing of lockdown and the same mobility restrictions over time.

We used strict inclusion and exclusion criteria to avoid mixing the effects of other immunosuppressive treatments in mortality. They were also useful to find an adequate patient profile who, a priori, should beneficiate of an anti-inflammatory treatment as corticosteroid pulses. We selected patients, at the inclusion time, with the onset of an acute respiratory distress syndrome.

To limit potential biases, we performed a propensity score matching (PSM) using the exact method. Thus, we obtained a more homogeneous sample with baseline features that were similar in both groups. Both exposed and non-exposed cohorts in the propensity score matching group had comparable Age, a similar pretest probability of dying (through the COVID-gram score), probability of being treated with corticosteroids, and uniform comorbidities. We again found in this PSM group, the same mortality decrease in the corticosteroid pulses arm. Then, to adjust other possible mortality causes, we performed a Cox regression multivariate model on the PSM group, once more finding a significant protective role of corticosteroid pulses in mortality.

Altogether, joining this strict inclusion and exclusión criteria with all this selection process set a reliable frame to compare mortality in both exposed and non-exposed groups. Thus, corticosteroid pulses might be a good option for the treatment of severe COVID-19, as they showed effective in reducing mortality in our cohort, and they are inexpensive and highly available worldwide. Our results can only extrapolate to patients with severe COVID-19, with a pO2/FiO2 lower than 300, not exposed to any kind of immunosuppression, and in the absence of a terminally ill situation at admission.

Some recent studies have confirmed that oral or intravenous low dose corticosteroids positively affect mortality (15). There was a decrease in mortality between 3·1 and 12·1 % (in the ICU admitted group), lower than the 12·6% of global mortality reduction (even higher in the ICU subset) that we found. We hypothesized that the effect of corticosteroid pulses might be higher than the low dose corticosteroids because they act in different pathways (genomic vs. Non-genomic) and behave, in fact, as different drugs (22). Our study is not powered to compare both low dose and pulse corticosteroid treatment, so we can not assure this statement. Future studies must investigate this topic.

Pulse corticosteroids did not reduce the ICU admission rate in our study. Most patients moved to ICU in the first 24 hours of hospitalization (83·6%), so we understand that those patients were critically ill at admission time, requiring ICU in any case.

The in-hospital stay was significantly longer in the pulses corticosteroid arm, but that differences disappear in the survivor’s group. Thus, this difference in the hospital stay was due to higher survival in the corticosteroid pulses group

The rate of adverse events and serious adverse events declared was similar in both groups. In the same way, in-hospital infection and viral shedding time were similar in both groups, but some of these adverse events and infections might be under-reported. The study was not powered to detect these adverse events because they were not always reported in the medical record in all patients.

In conclusion, this study provides evidence about the treatment with corticosteroid pulses in severe COVID-19 that might significantly reduce mortality. This data must be confirmed in prospective randomized studies.

## Data Availability

We can provide all data in the study if requested with a funded reason.

## ABBREVIATIONS

ARDS: Acute respiratory distress syndrome
COVID-19: Coronavirus disease 2019
ICU: Intensive care unit
PSM: Propensity score matching
SARS-CoV-2: Severe acute respiratory syndrome coronavirus 2
PCR: Polymerase chain reaction
IFN—γ: Interferon-γ
TNF-α: Tumor necrosis factor α
LDH: Lactate dehydrogenase
CRP: C-reactive protein

## BULLET POINTS

- Corticosteroid pulses can improve mortality in severe COVID-19 patients
- The Kaplan Meier curve with the propensity score matching and multivariate analysis in this group is reliable
- There are no substantial changes in ICU admission and hospital stay

