## supplementary material for "“Corticosteroid pulses for hospitalized patients with COVID-19: Effects on mortality”"

### SECTION A: VARIABLE DESCRIPTION AND CODING IN THE STUDY:

- **Variables related to mortality:**
  - X-Ray impairment
  - Age
  - Hemopyisis
  - Low consciousness level
  - SaO2 at inclusion
  - FiO2 at inclusion
  - Cancer
  - Neutrophil / Lymphocyte index
  - LDH
  - Direct Bilirubin
  - Number of comorbidities from 1 to 4 from these: chronic
  - obstructive pulmonary disease, diabetes, hypertension, coronary
  - artery disease, cerebrovascular disease, hepatitis B, cancer,
  - chronic renal disease, immunodeficiency disease, and
  - pregnancy.
  - Modified Charlson index (Age, coronary artery disease, chronic heart failure, peripheral vascular disease, cerebrovascular disease, dementia, chronic obstructive pulmonary disease, connective tissue disease, peptic ulcer, liver disease, chronic kidney disease, diabetes, solid tumor, hemiplegia, leukemia, lymphoma, AIDS).
  - COVID-GRAM calculation
  - Sex
  - Glucose
  - Procalcitonin
  - CRP
  - IL-6
  - D-Dimer
  - Creatinin
  - Ferritin
  - Obese (Y/N)
  - Epidemiologic week (week zero matches with the first death in Spain, March 3rd)
  - Date of symptoms onset
  - Stroke during hospitalization
  - Myocardial infarction during hospitalization
  - Pulmonary embolism during hospitalization
  - Procalcitonin > 0.5 at any time
  - Microbiological isolation (Y/N) and describe the isolation.
  - Confirmed in-hospital infection (Y/N)
  - Peak LDH
  - Lowest Lymphocyte count
  - Peak D-Dimer
  - Peak ferritin
  - Peak CRP

  * Blood test data must be referred to the blood test in the ±3 days in which patients reach de inclusion criterion.

- **Concomitant treatments: (not allowed immunosuppressors)**

  - Hydroxychloroquine

- Azithromycin
- Interferon-beta-1-b
- Lopinavir / Ritonavir
- Remdesivir

- **Inclusion / Exclusion criteria assessment:**
  - Date when PaFi decrease under 300, directly or indirectly measured:
    - PaO2
    - FiO2
    - Pulse oxymetry when Pa/Fi decreased under 300 (estimated by non-linear imputation of SaFi to PaFi).

      * PaFi <300 must be sustained more than 24 hours or repeated at least three times on different days. An isolated low value is not considered for inclusion.

      * Values can be measured in the emergency department or during the hospitalization.
  - Pregnancy
  - Age under 18

  - Death in the first 24 hours: Admission date and death date
  - Date of first SARS-Cov-2 PCR on the nasopharyngeal swab (or date of positive Elisa serology if compatible clinical features)
  - Treatment with classic immunosuppressors (Y/N)
  - Treatment with cytokine blockers like tocilizumab or anakinra (Y/N)
  - Limitation of therapeutic effort or terminally ill patient at the admission days or within the first 24 hours of hospitalization (Y/N).
  - On treatment with corticosteroids in a different way than described in inclusión criteria (Y/N)

- **Outcome related variables:**
  - Admission date
  - Discharge date
  - Date of death (if applicable). Register if the patient died at 30 days once he was discharged, (Y/N).
  - If the patient died: Cardiovascular death (Y/N). In the non-cardiovascular death subgroup register: Shock (1), refractory hypoxemia (3)0, multiorgan failure (3), Other (4). In the cardiovascular death subgroup register: Myocardial infarction(1), Arrhythmia (2), heart failure (3), cardiogenic shock (4), Other (5)
  - Date of first negative SARS-Cov-2 PCR.
  - Reach the definition of superinfection: PCT > 0.5
  - ICU admission date
  - The need for ventilatory support (invasive ventilation 1, non-invasive ventilation 2, no assistance needed 0)

- **Corticosteroid exposure:**

  * Pulse corticosteroids are defined as 125-500 mg daily, for two to five days, within the ±3 days in which patients reach the inclusion criterion.
  - Date onset of the first corticosteroid pulse
  - The second course of corticosteroid pulses (Y/N)

### SECTION B: CONSTRUCTION OF THE PROPENSITY SCORE AND MATCHING PROCESS:

The variables that have a statistical association in the univariate analysis between treatment with corticosteroid boluses (dichotomous 0 or 1) were the following:

- Center of Castilla y León (Hospital Clínico Universitario de Valladolid, Hospital Universitario de Salamanca, and Hospital Universitario de Burgos)
- The epidemiological week from the first positive PCR in early March
- Classic Charlson index
- Treatment with lopinavir/ritonavir at admission
- Treatment with azithromycin at admission
- Treatment with Interferon beta 1b at admission
- Treatment with colchicine on admission
- Need for non-invasive mechanical ventilation
- Presence of unilateral or bilateral radiological infiltrates
- Glucose at admission in mg/dl
- Ferritin at admission
- Ferritin or at admission or a week
- D-Dimer upon admission
- CRP upon admission
- D-Dimer upon admission or in the first week
- Dichotomous Charlson index (up to 2, and 3 or more comorbidities)

There is **no** statistical association between being treated with corticosteroids and the following variables:

- Age
- COVID-GRAM score (p-value between 0.05 and 0.1)
- Modified Charlson index
- Sex
- Hypertension
- Type 2 diabetes
- Dyslipidemia
- Smoker
- Obese
- Chronic ischemic heart disease

- Chronic kidney disease
- Significant lung disease
- History of cancer
- HIV
- Cerebrovascular disease
- Hemiplegia
- Previous rheumatological disease
- Moderate liver disease
- Presence of cirrhosis
- Peptic ulcer
- Liver disease of any severity
- Heart failure
- Dementia
- Peripheral artery disease
- Hemoptysis
- Low level of consciousness
- Previous treatment with oral anticoagulants
- Need for admission to ICU
- Need for invasive mechanical ventilation
- Presence of respiratory failure at the time of admission
- Degree of anticoagulation during admission
- Complications in the form of Pulmonary embolism, stroke, hemorrhage, or heart attack
- Neutrophils at admission and in the first week
- Lymphocytes at admission and in the first week
- Creatinine at admission and in the first week
- LDH at admission and in the first week
- CPK at admission and per week
- IL-6 levels both upon admission and the first week
- Direct bilirubin on admission
- Ferritin a week after admission
- D-Dimer one week after admission
- CRP and procalcitonin per week
- Maximum values of LDH, ferritin, D-dimer, and CRP
- Minimum lymphocyte values

- Time from the onset of symptoms to admission

- Time from the onset of symptoms to PaFi less than 300

**Selected variables were dichotomized**, and the cutoff points selected for the non-dichotomous variables were the following:

- Blood glucose 162 mg / dl

- We opted for the dichotomized Charlson with a score or <2 or 2 or more than 2.

- Ferritin on admission or per week: 633

- CRP upon admission 95.55

- D-Dimer upon admission or per week: 1005

The qualitative variables of more than 2 categories are made dichotomous in this way:

Center: Hospital Clínico Universitario de Valladolid versus Hospital Universitario de Burgos or Hospital Universitario de Salamanca.

Epidemiological week: One group from weeks 0 to 2, and the other from week 3 onwards.

**Raw OR when doing binary logistic regression:**

Dichotomous epidemiological week: OR 3.8

Dichotomous center: OR 0.48

Glucose NS (neither cutoff point 160 nor 105 mg/dl)

Ferritin cutoff 633: OR 2.65

PCR cut 65.5: OR 2.28

D-dimer cut 1005 and cut 575: NS

Dichotomous Charlson: OR 0.5

Lopinavir/ritonavir treatment: 2.58

Azithromycin treatment: OR 5.22

Interferon beta 1b treatment: OR 3.12

Infiltrates in the Rx: OR 2.36

**The variables that remain in the final multivariate model are:**

Epidemiological week

Bilateral infiltrates or not

Center

Ferritin

Covid gram score

The final model has an area under the curve of  73.7%.

The distribution of the propensity score between those exposed and the not exposed ones to corticosteroids is this:

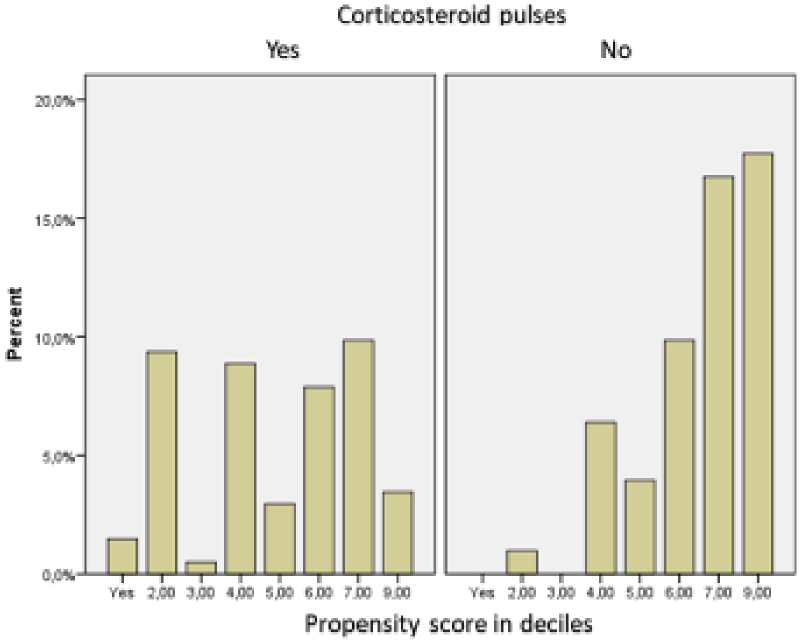


We propose 7 propensity score matching models using various mathematical methods. We compared the 7 models that a priori were better, by the following methods:

Nearest neighbor

Exact matching

Nearest neighbor con caliper de 0.05

Nearest neighbor con caliper de 0.1

Nearest neighbor con caliper de 0.2

Nearest neighbor con caliper de 0.25

Nearest neighbor con caliper de 0.30

We describe the results of the six methods used for preprocessing

### 1. Nearest neighbor method:

```
Summary of balance for all data:
                          Means Treated Means Control SD Control Mean Diff eQQ Med eQQ Mean eQQ M
ax
distance                         0.6020        0.4151     0.2220    0.1869     0.2   0.1849  0.33
53
semana_epidem_dicotom            0.8843        0.6983     0.4610    0.1860     0.0   0.1810  1.00
00
centro_dicotom                   0.3306        0.6466     0.4801   -0.3160     0.0   0.3190  1.00
00
Rx_infiltrado_dicotom            0.8264        0.6724     0.4714    0.1540     0.0   0.1466  1.00
00
ferrit633realyestimada           0.7025        0.5000     0.5022    0.2025     0.0   0.1983  1.00
00
covidgram123                     0.8347        0.7500     0.4349    0.0847     0.0   0.0776  1.00
00


Summary of balance for matched data:
                          Means Treated Means Control SD Control Mean Diff eQQ Med eQQ Mean eQQ M
ax
distance                         0.6167        0.4151     0.2220    0.2016  0.2153   0.2016  0.34
28
semana_epidem_dicotom            0.8879        0.6983     0.4610    0.1897  0.0000   0.1897  1.00
00
centro_dicotom                   0.3017        0.6466     0.4801   -0.3448  0.0000   0.3448  1.00
00
Rx_infiltrado_dicotom            0.8534        0.6724     0.4714    0.1810  0.0000   0.1810  1.00
00
ferrit633realyestimada           0.7069        0.5000     0.5022    0.2069  0.0000   0.2069  1.00
00
covidgram123                     0.8448        0.7500     0.4349    0.0948  0.0000   0.0948  1.00
00

Percent Balance Improvement:
                          Mean Diff. eQQ Med eQQ Mean eQQ Max
distance                     -7.8531 -7.6163  -9.0251 -2.2358
semana_epidem_dicotom        -1.9533  0.0000  -4.7619  0.0000
centro_dicotom               -9.1319  0.0000  -8.1081  0.0000
Rx_infiltrado_dicotom       -17.5301  0.0000 -23.5294  0.0000
ferrit633realyestimada       -2.1816  0.0000  -4.3478  0.0000
covidgram123                -11.9428  0.0000 -22.2222  0.0000

Sample sizes:
          Control Treated
All           116     121
Matched       116     116
Unmatched       0       5
Discarded       0       0
```

```
>  m.out <- matchit(bolos_corticoides ~ semana_epidem_dicotom + centro_dicotom + Rx_infiltrado
_dicotom + ferrit633realyestimada + covidgram123, data=boloscortis, method= "nearest")
Warning message:
In matchit2nearest(c(`1` = 0, `2` = 0, `3` = 0, `4` = 0, `5` = 0,  :
  Fewer control than treated units and matching without replacement.  Not all treated units wi
ll receive a match.  Treated units will be matched in the order specified by m.order: largest
> bal.tab(m.out)
Call
 matchit(formula = bolos_corticoides ~ semana_epidem_dicotom + centro_dicotom + Rx_infiltrado_
dicotom + ferrit633realyestimada + covidgram123, data = boloscortis, method = "nearest")

Balance Measures
                           Type Diff.Adj
distance               Distance   0.9083
semana_epidem_dicotom    Binary   0.1897
centro_dicotom           Binary  -0.3448
Rx_infiltrado_dicotom    Binary   0.1810
ferrit633realyestimada   Binary   0.2069
covidgram123             Binary   0.0948

Sample sizes
          Control Treated
All           116     121
Matched       116     116
Unmatched       0       5
>
```

**Love plot of the model:**

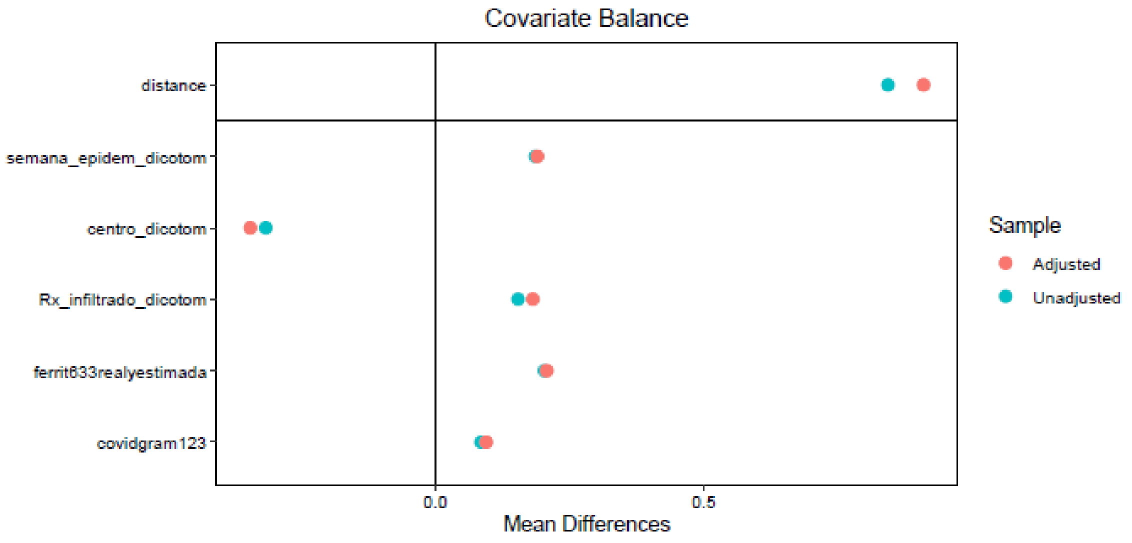


**Distribution of propensity scores in the model before and after matching:**

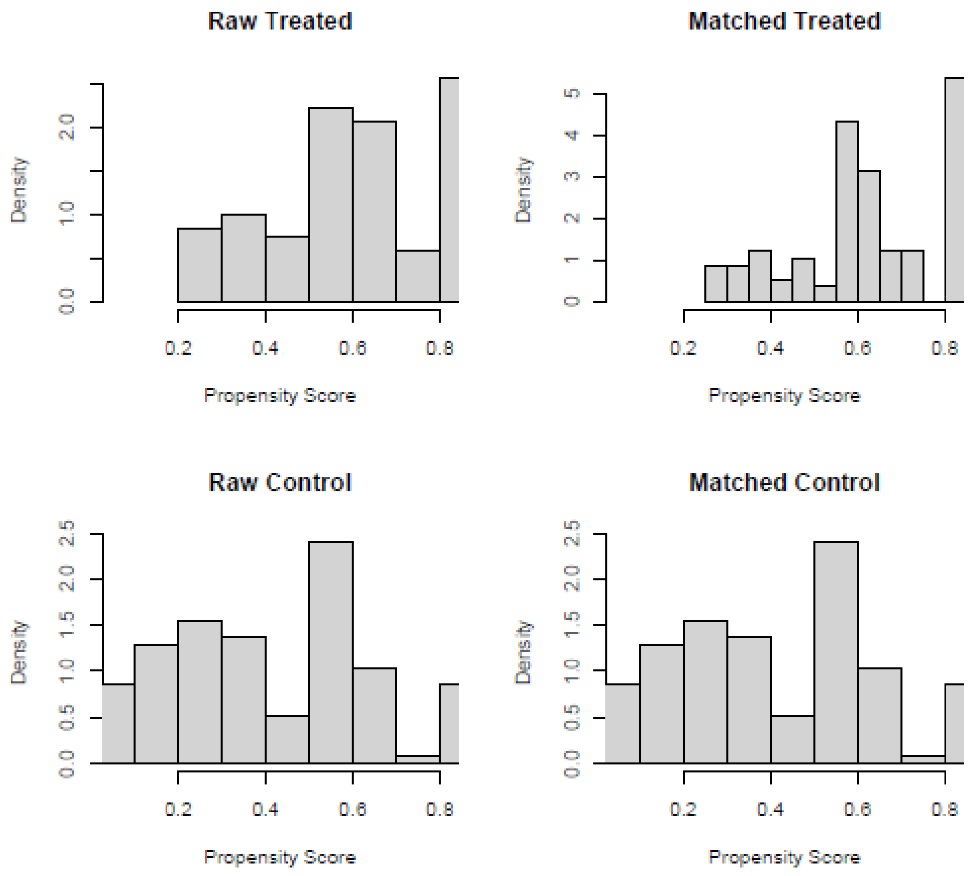

### 2. Exact matching method:

```
Call:
matchit(formula = bolos_corticoides ~ semana_epidem_dicotom +
    centro_dicotom + Rx_infiltrado_dicotom + ferrit633realyestimada +
    covidgram123, data =boloscortis, method = "exact")

Sample sizes:
          Control Treated
All           116     121
Matched        88     119
Discarded      28       2

Matched sample sizes by subclass:
   Treated Control Total
1        2       1     3
2        2       5     7
3        2       2     4
4        2       3     5
5        7       1     8
6        3       3     6
7        4       2     6
8        2       7     9
9        3       1     4
10      18       9    27
11       5      11    16
12       7       3    10
13       2       3     5
14       3       5     8
15      31      10    41
16       6       6    12
17      20      16    36
```

```
> bal.tab(m.out)
Note: 's.d.denom' not specified; assuming pooled.
Call
 matchit(formula = bolos_corticoides ~ semana_epidem_dicotom + centro_dicotom + Rx_infiltrado_
dicotom + ferrit633realyestimada + covidgram123, data = boloscortis, method = "exact")

Balance Measures
                         Type Diff.Adj
semana_epidem_dicotom  Binary        0
centro_dicotom         Binary        0
Rx_infiltrado_dicotom  Binary        0
ferrit633realyestimada Binary       -0
covidgram123           Binary       -0

Sample sizes
                     Control Treated
All                  116.000     121
Matched (ESS)         53.938     119
Matched (Unweighted)  88.000     119
Unmatched             28.000       2
>
```

We do not show the raw and matched distribution because, in the exact method matching, all propensity scores are precisely the same in both groups.

**Love plot of the model:**

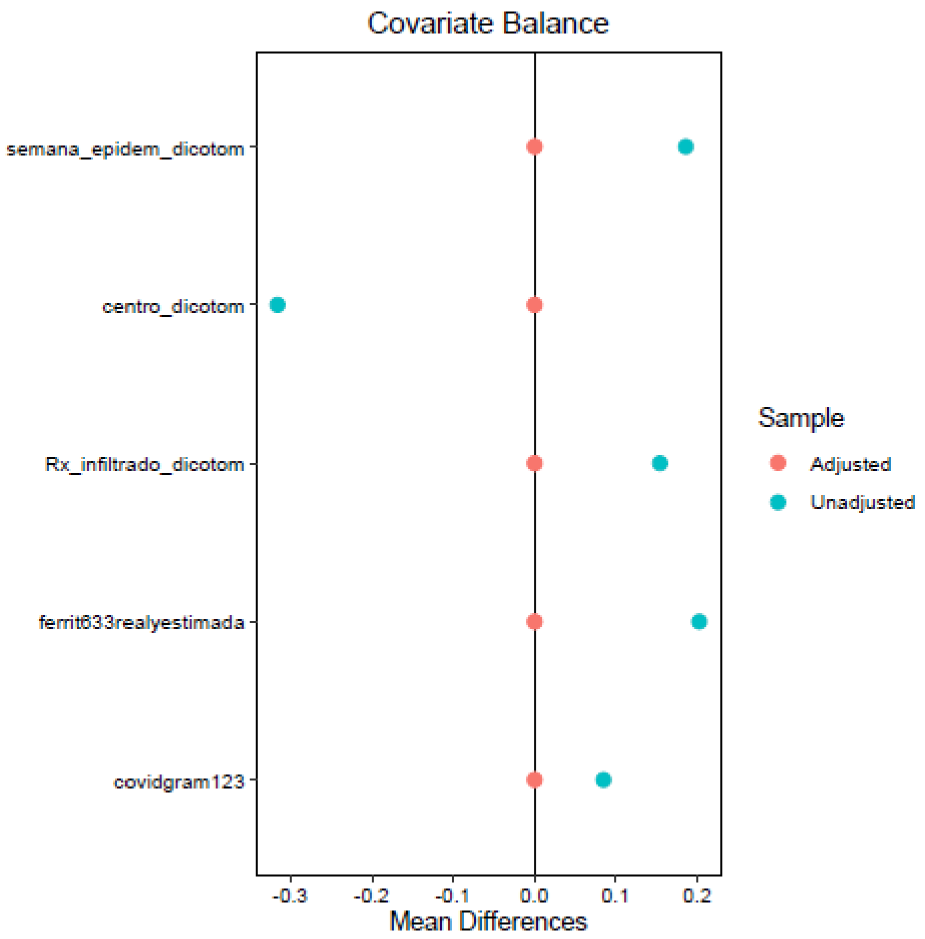


### 3. Nearest neighbor with a caliper at 0.05:

```
> bal.tab(m.out)
Note: 's.d.denom' not specified; assuming pooled.
Call
 matchit(formula = bolos_corticoides ~ semana_epidem_dicotom + centro_dicotom + Rx_infiltrado_
dicotom + ferrit633realyestimada + covidgram123, data = boloscortis, method = "nearest", calip
er = 0.05)


Balance Measures
                            Type Diff.Adj
distance                Distance  -0.0005
semana_epidem_dicotom     Binary   0.0143
centro_dicotom            Binary   0.0000
Rx_infiltrado_dicotom     Binary  -0.0143
ferrit633realyestimada    Binary  -0.0143
covidgram123              Binary   0.0143

Sample sizes
          Control Treated
All           116     121
Matched        70      70
Unmatched      46      51
>
```

**Distribution of propensity scores in the model before and after matching:**

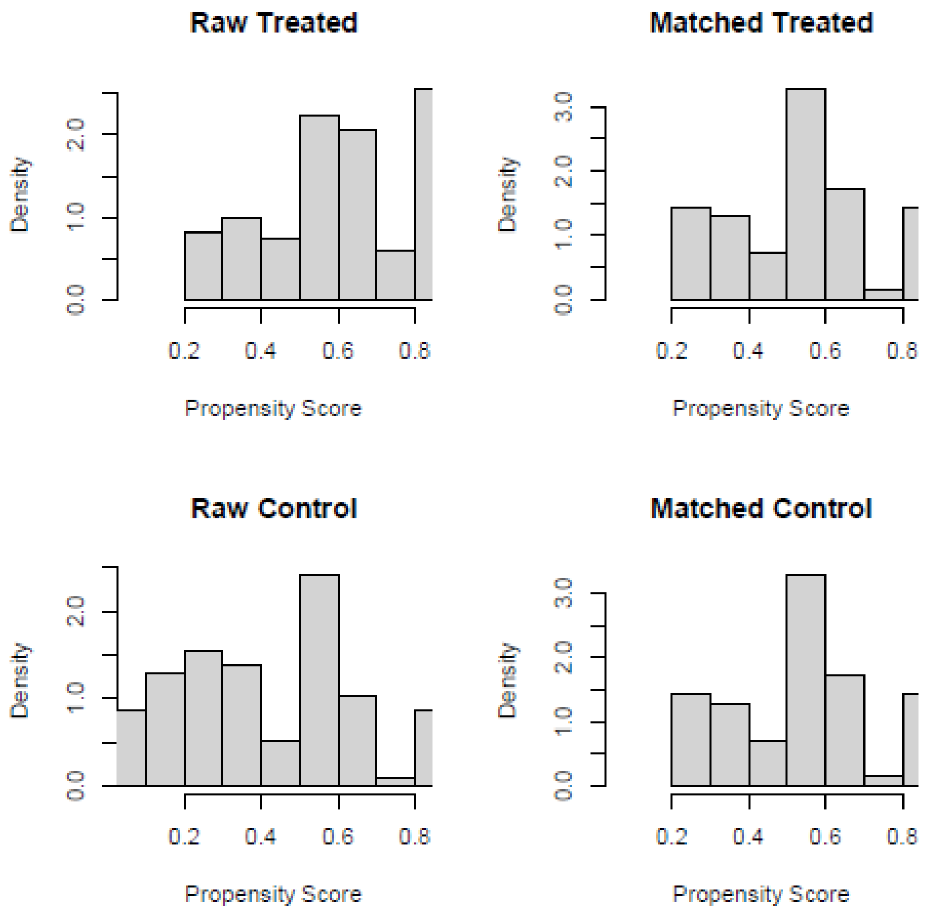


**Love plot of the model:**

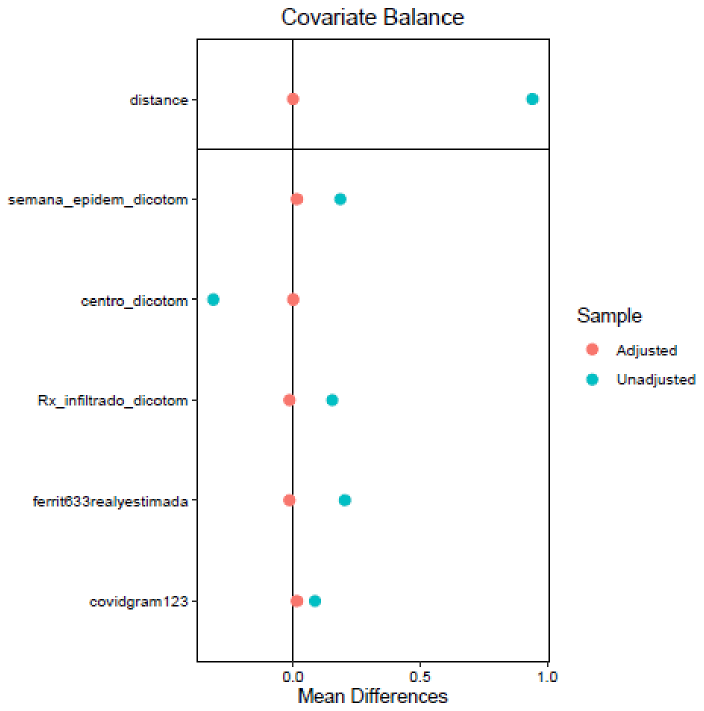

#### 4. Nearest neighbor with a caliper at 0.1:

```
> bal.tab(m.out)
Note: 's.d.denom' not specified; assuming pooled.
Call
 matchit(formula = bolos_corticoides ~ semana_epidem_dicotom + centro_dicotom + Rx_infiltrado_
dicotom + ferrit633realyestimada + covidgram123, data = boloscortis, method = "nearest", calip
er = 0.1)

Balance Measures
                            Type Diff.Adj
distance                Distance   0.0011
semana_epidem_dicotom     Binary  -0.0143
centro_dicotom            Binary   0.0143
Rx_infiltrado_dicotom     Binary   0.0429
ferrit633realyestimada    Binary   0.0143

covidgram123              Binary  -0.0143

Sample sizes
          Control Treated
All           116     121
Matched        70      70
Unmatched      46      51
>
```

**Distribution of propensity scores in the model before and after matching:**

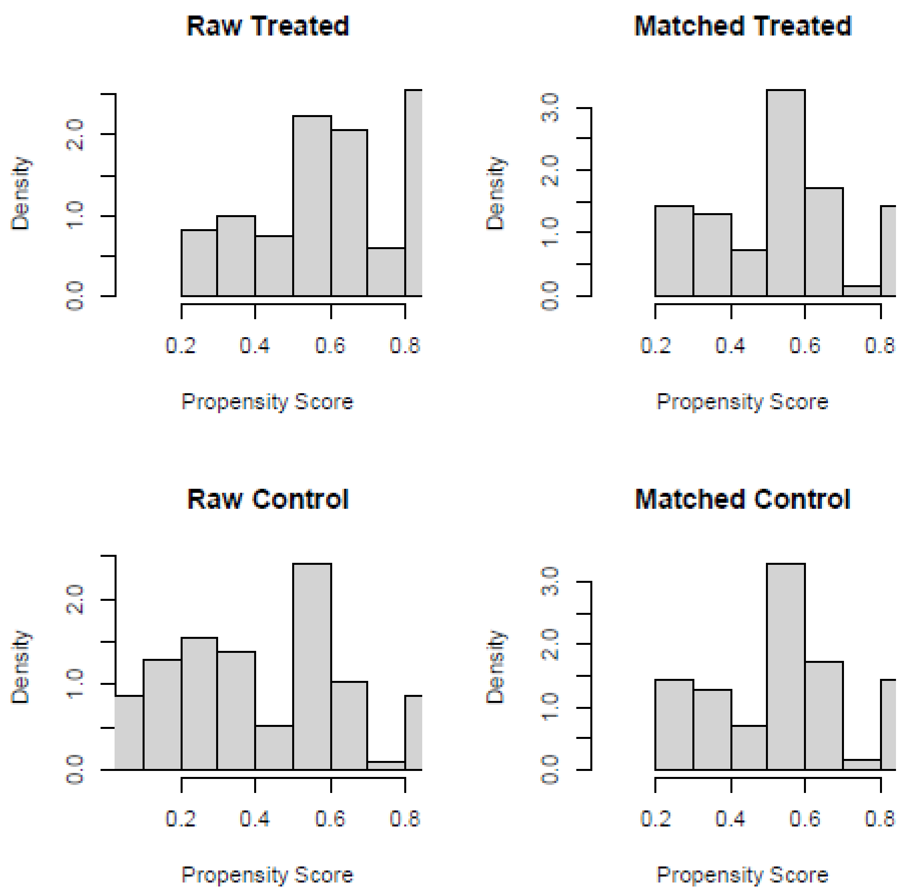

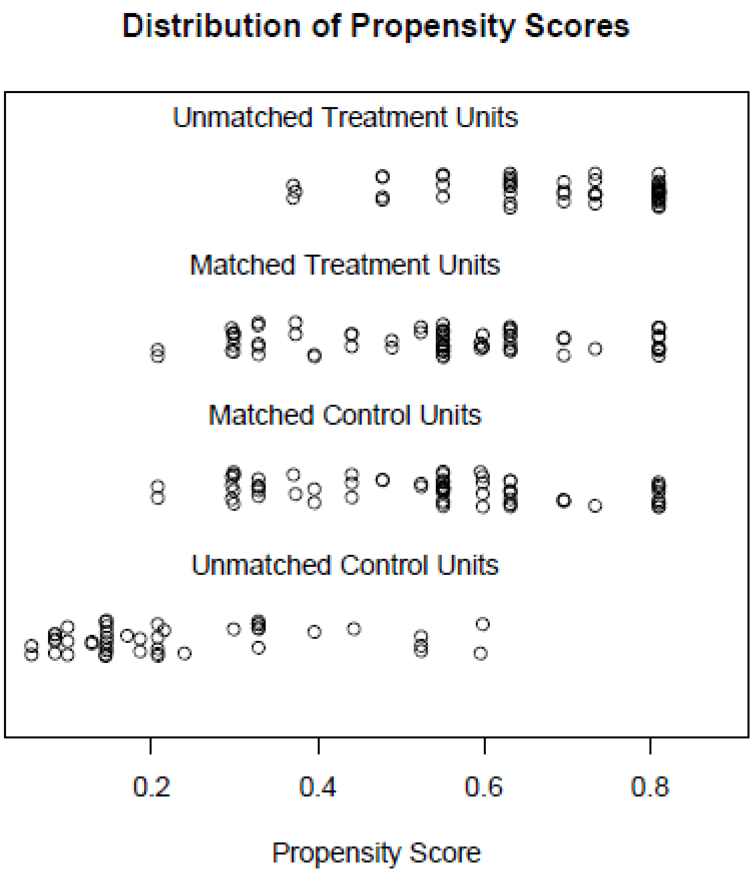


**Love plot of the model:**

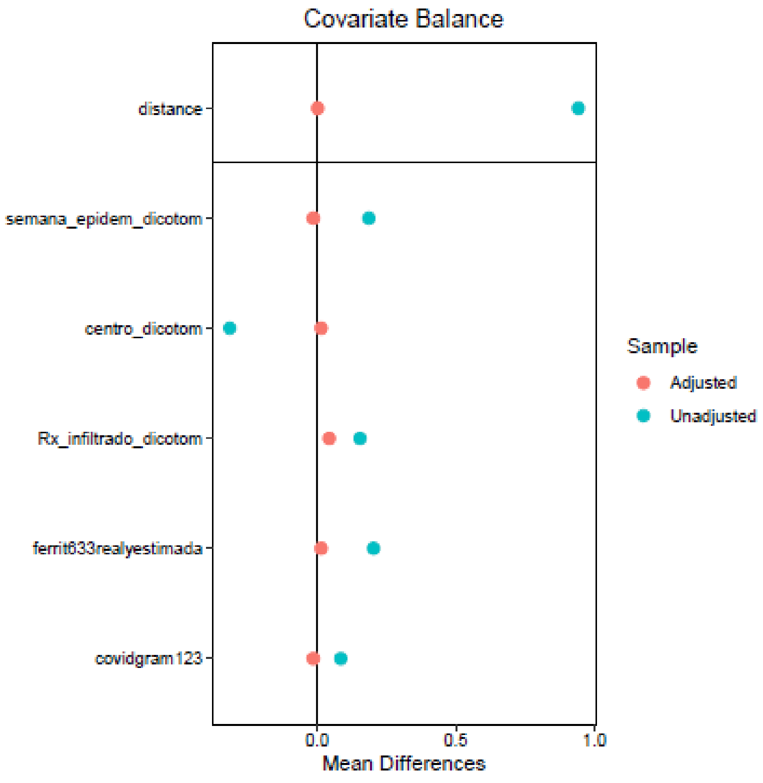

### 5. Nearest neighbor with a caliper at 0.2:

```
> bal.tab(m.out)
Note: 's.d.denom' not specified; assuming pooled.
Call
 matchit(formula = bolos_corticoides ~ semana_epidem_dicotom + centro_dicotom + Rx_infiltrado_
dicotom + ferrit633realyestimada + covidgram123, data = boloscortis, method = "nearest", calip
er = 0.2)

Balance Measures
                           Type Diff.Adj
distance               Distance   0.0381
semana_epidem_dicotom    Binary  -0.0127
centro_dicotom           Binary  -0.0253
Rx_infiltrado_dicotom    Binary   0.0506
ferrit633realyestimada   Binary  -0.0380
covidgram123             Binary   0.0380

Sample sizes
          Control Treated
All           116     121
Matched        79      79
Unmatched      37      42
>
```

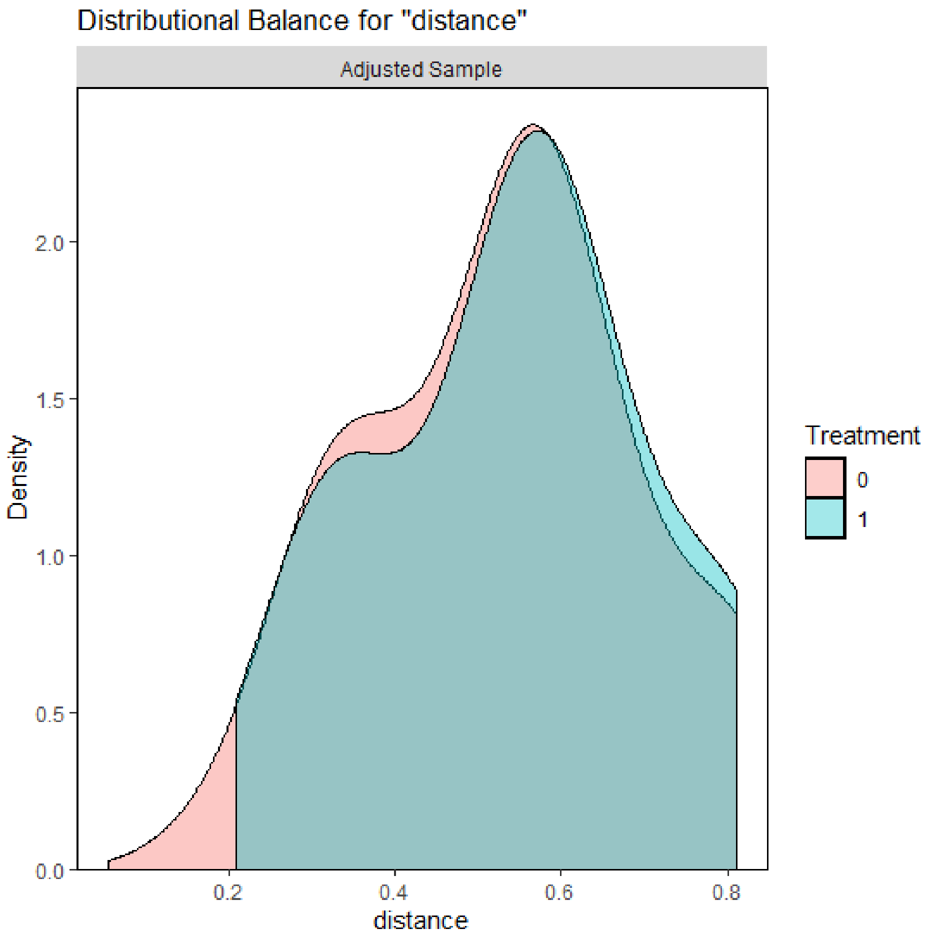

**Distribution of propensity scores in the model before and after matching:**

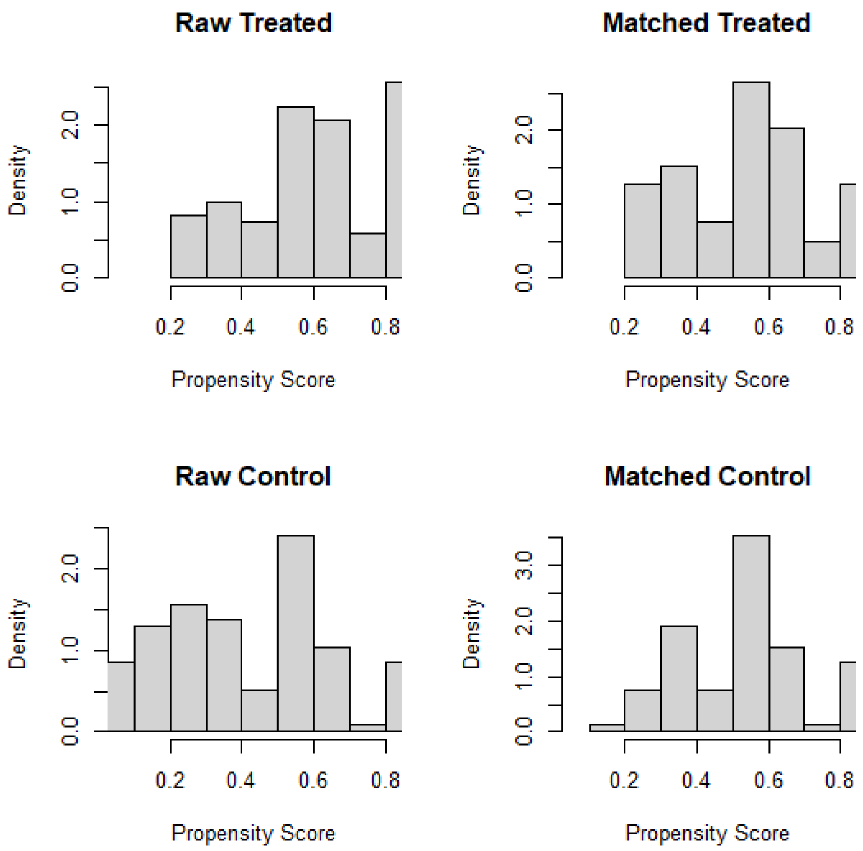


**Love plot of the model:**

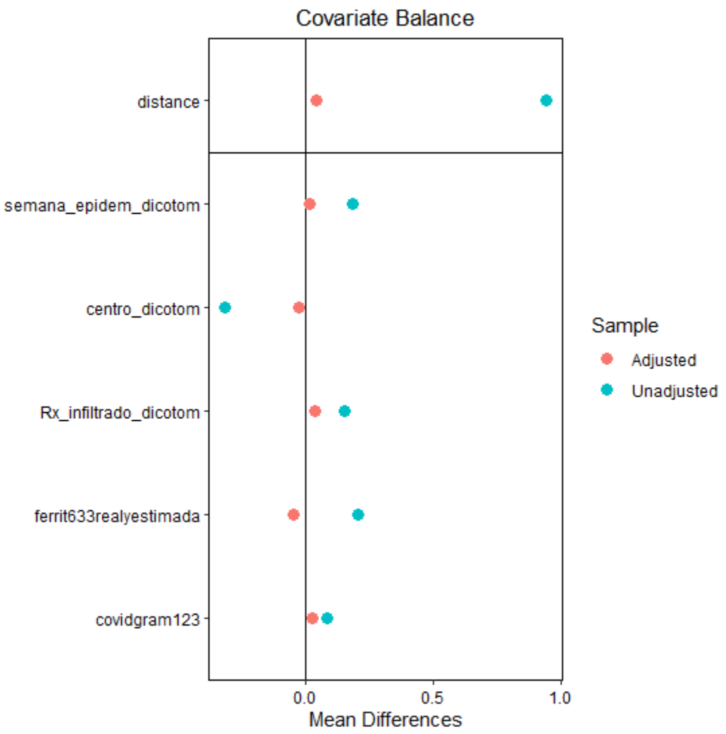

#### 6. Nearest neighbor with a caliper at 0.25:

```
> bal.tab(m.out)
Note: 's.d.denom' not specified; assuming pooled.
Call
 matchit(formula = bolos_corticoides ~ semana_epidem_dicotom + centro_dicotom + Rx_infiltrado_
dicotom + ferrit633realyestimada + covidgram123, data = boloscortis, method = "nearest", calip
er = 0.25)

Balance Measures
                            Type Diff.Adj
distance                Distance   0.0844
semana_epidem_dicotom     Binary  -0.0122
centro_dicotom            Binary  -0.0732
Rx_infiltrado_dicotom     Binary   0.0488
ferrit633realyestimada    Binary  -0.0244
covidgram123              Binary  -0.0366

Sample sizes
          Control Treated
All           116     121
Matched        82      82
Unmatched      34      39
```

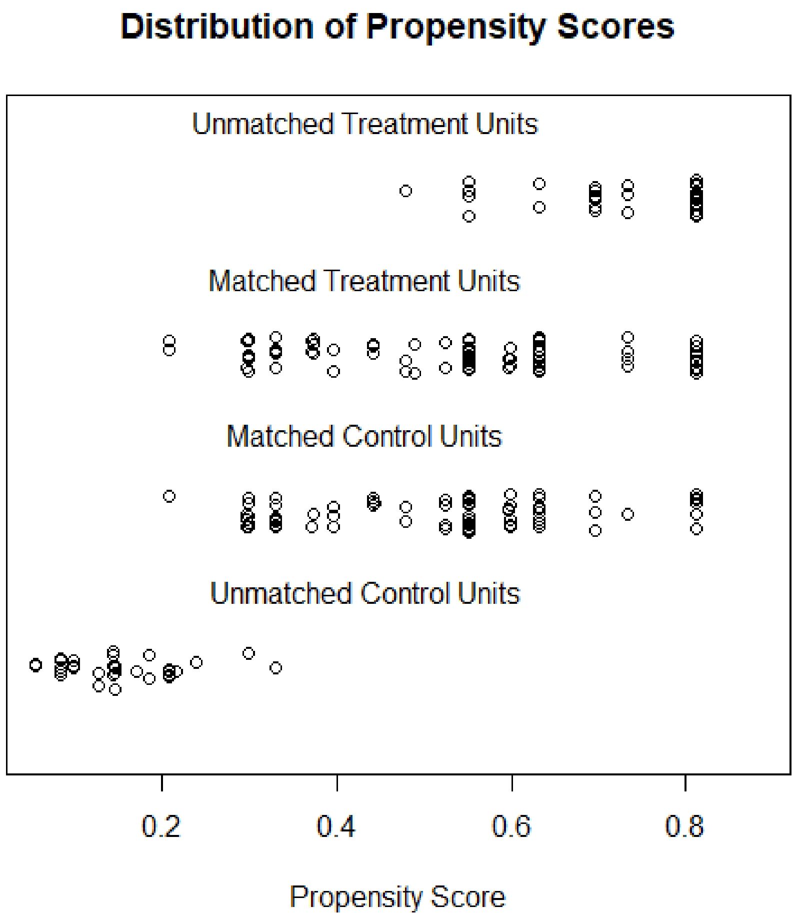

**Distribution of propensity scores in the model before and after matching:**

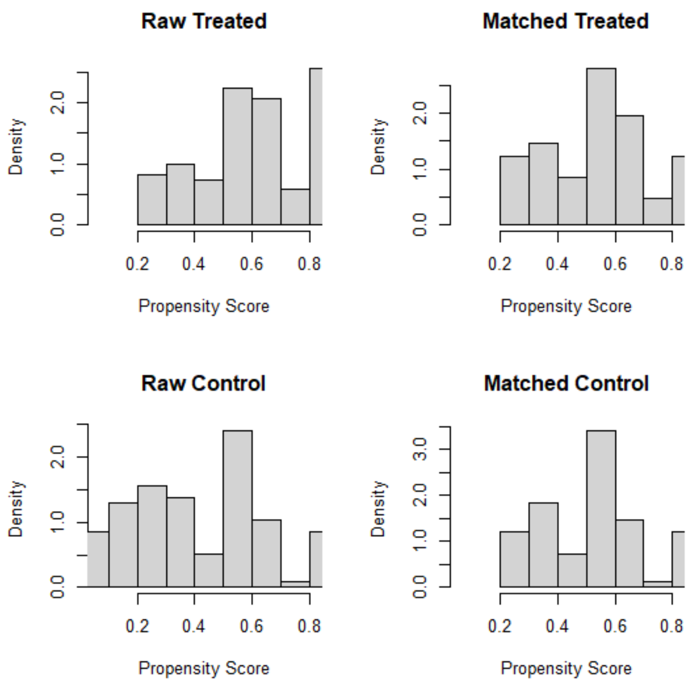


**Love plot of the model:**

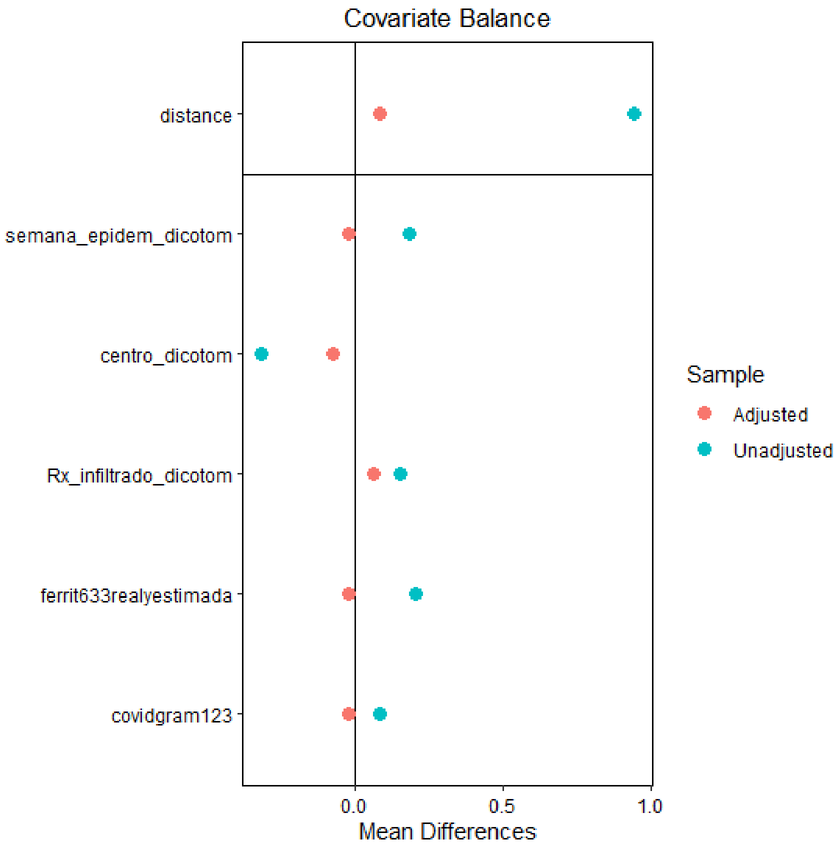

### 7. Nearest neighbor with caliper at 0.3:

```
> bal.tab(m.out)
Note: 's.d.denom' not specified; assuming pooled.
Call
 matchit(formula = bolos_corticoides ~ semana_epidem_dicotom + centro_dicotom + Rx_infiltrado_
dicotom + ferrit633realyestimada + covidgram123, data = boloscortis, method = "nearest", calip
er = 0.3)

Balance Measures
                           Type Diff.Adj
distance               Distance   0.1179
semana_epidem_dicotom    Binary  -0.0244
centro_dicotom           Binary  -0.0732
Rx_infiltrado_dicotom    Binary  -0.0244
ferrit633realyestimada   Binary   0.0610
covidgram123             Binary  -0.0122

Sample sizes
          Control Treated
All           116     121
Matched        82      82
Unmatched      34      39
>
```

**Distribution of propensity scores in the model before and after matching:**

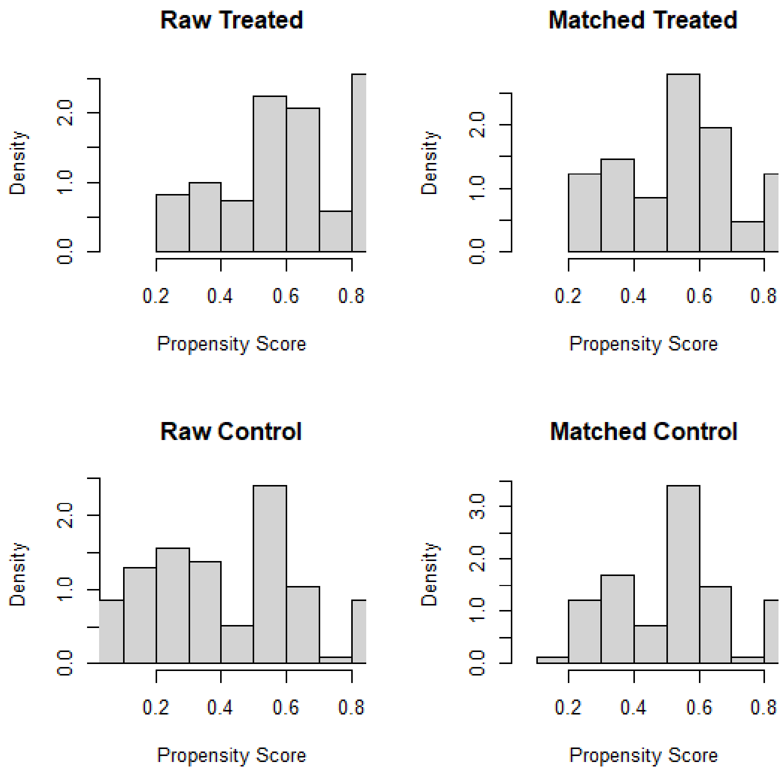

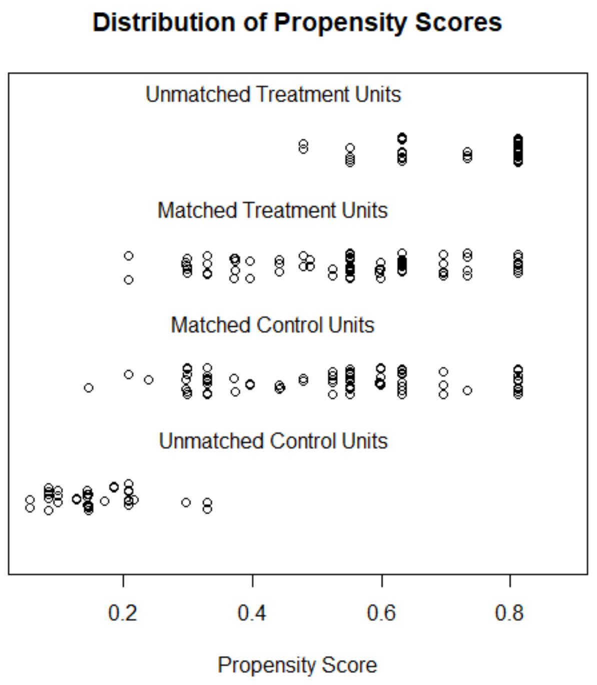


**Love plot of the model:**

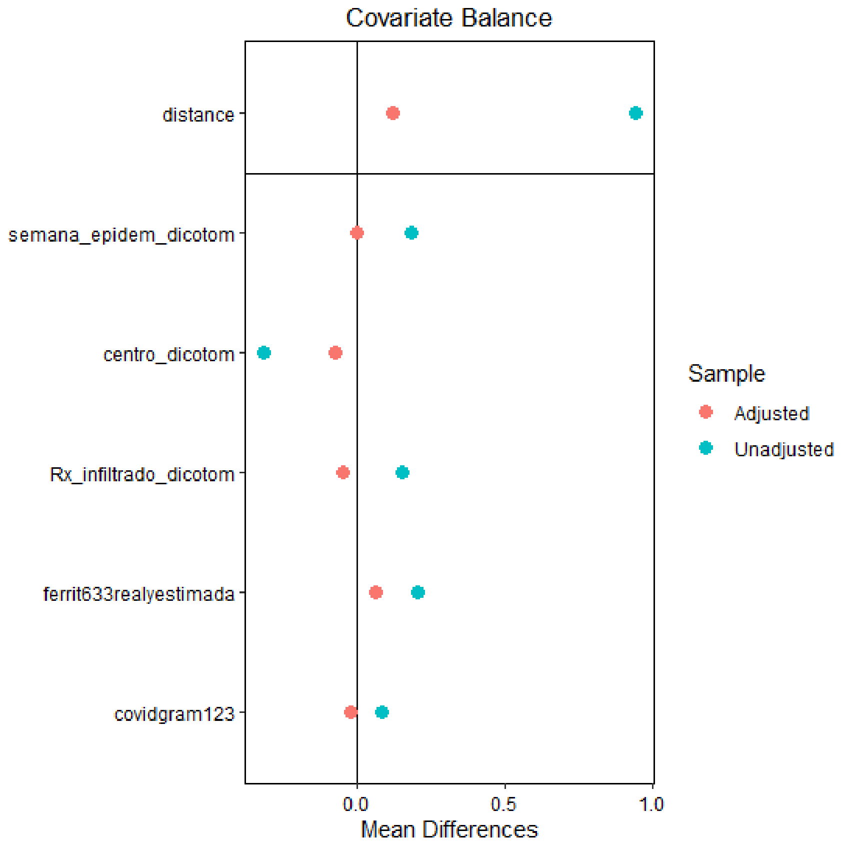

We chose the **exact matching method** from all the available models because that is the one that loses the least number of patients with a better adjustment in terms of distance (propensity score) in those treated respecting controls.

**Secondary endpoints:**

**30-days mortality:** Kaplan Meier Curve in all participants and the propensity score matching group.

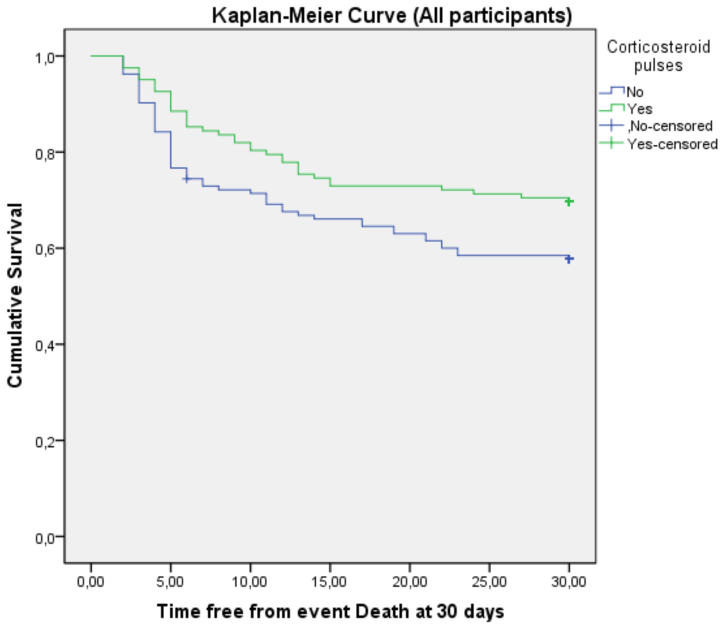


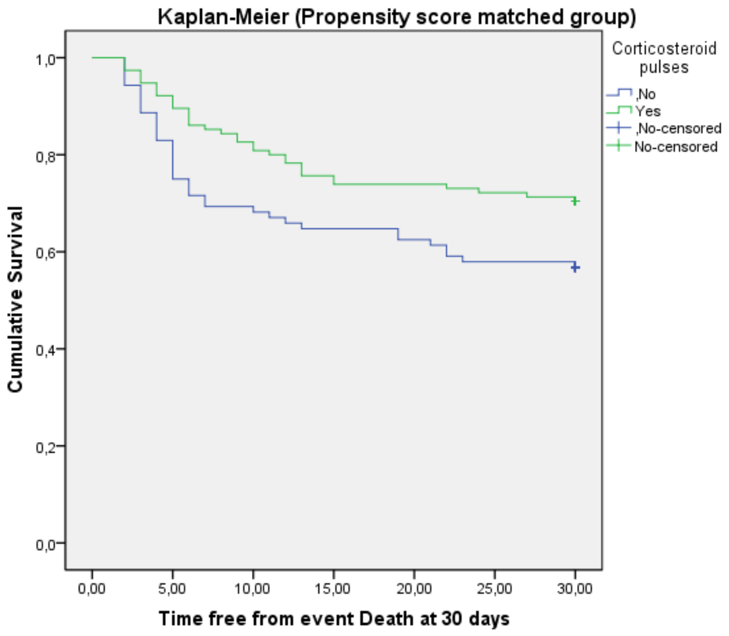


Cox regression model to predict 30-days mortality in the propensity score-matched group:

**Variables in the Equation**

| | B | SE | Wald | df | p-value | OR | 95,0% CI for OR | |
|---|---|---|---|---|---|---|---|---|
| | | | | | | | Lower | Upper |
| Corticosteroid pulses | -,554 | ,281 | 3,877 | 1 | ,049 | ,575 | ,331 | ,997 |
| Age higher than 80 years | 2,048 | ,317 | 41,614 | 1 | ,000 | 7,753 | 4,161 | 14,445 |
| Neutrophil (Lymphocyte index higher than 7.4 | ,837 | ,294 | 8,117 | 1 | ,004 | 2,309 | 1,298 | 4,105 |
| Peak LDH > 372 UI/ml | ,726 | ,300 | 5,844 | 1 | ,016 | 2,067 | 1,147 | 3,725 |
| CRP > 200 mg/dl | 1,254 | ,309 | 16,515 | 1 | ,000 | 3,505 | 1,914 | 6,417 |
| Charlson index with 3 or more comorbidities | ,700 | ,300 | 5,441 | 1 | ,020 | 2,014 | 1,118 | 3,628 |

### Survival in ICU-admitted patients:

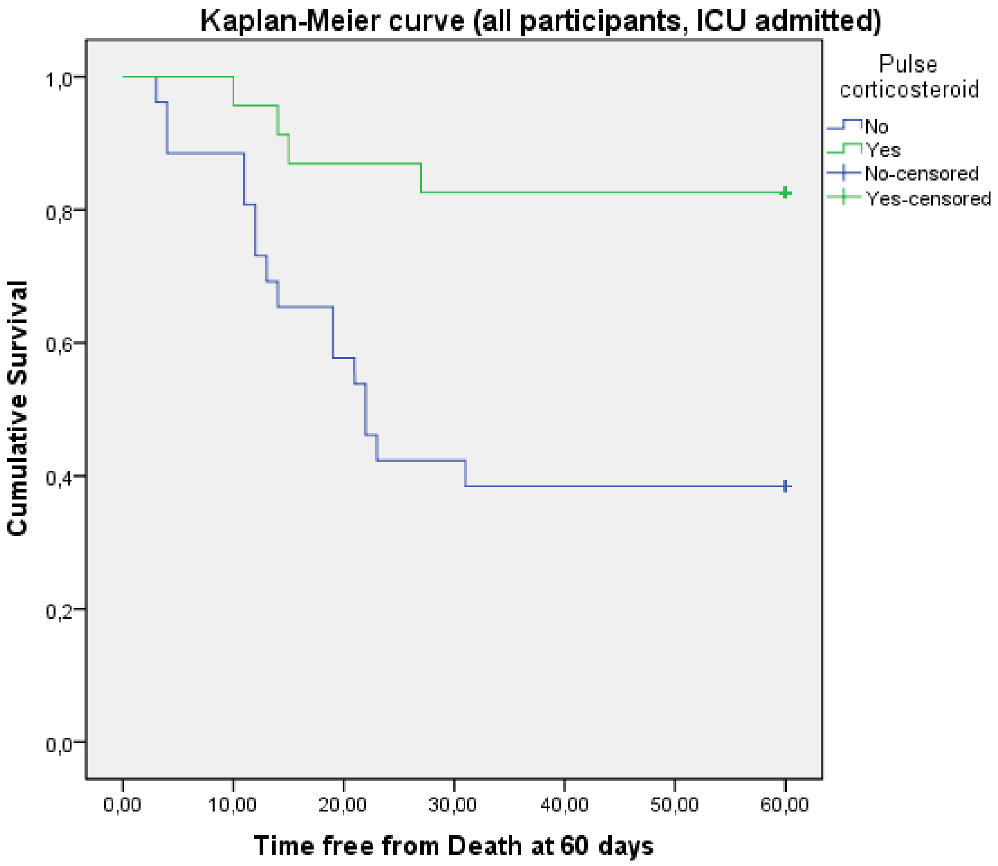

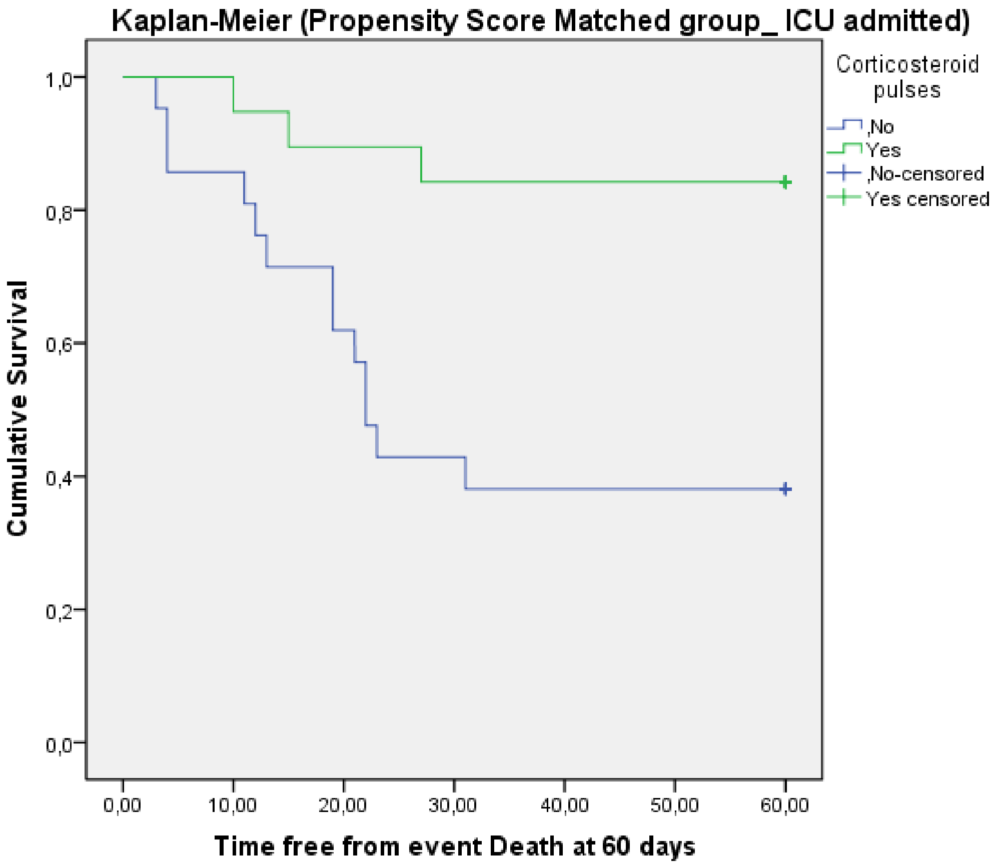


After adjusting variables in a Cox regression, we cannot make sure that corticosteroid pulses reduce mortality:

**Variables in the Equation**

| | B | SE | Wald | df | p-value | OR | 95,0% CI for Exp(B) | |
|---|---|---|---|---|---|---|---|---|
| | | | | | | | Lower | Upper |
| Corticosteroid pulses | -,405 | ,788 | ,264 | 1 | ,607 | ,667 | ,142 | 3,124 |
| D-Dimer >1621 U/ml | 4,468 | 1,206 | 13,727 | 1 | ,000 | 87,198 | 8,203 | 926,906 |
| SaFi >217 | 1,995 | ,930 | 4,603 | 1 | ,032 | 7,354 | 1,188 | 45,516 |
| Peak LDH >564 | 2,884 | 1,092 | 6,979 | 1 | ,008 | 17,894 | 2,105 | 152,101 |
| Peak CRP >257 mg/dl | 1,989 | ,906 | 4,819 | 1 | ,028 | 7,310 | 1,238 | 43,178 |
| Age>68 | 2,348 | ,884 | 7,051 | 1 | ,008 | 10,465 | 1,850 | 59,211 |
| More than 1 comorbiditie | 2,727 | ,870 | 9,827 | 1 | ,002 | 15,284 | 2,779 | 84,073 |
